# Accelerated SARS-CoV-2 intrahost evolution leading to distinct genotypes during chronic infection

**DOI:** 10.1101/2022.06.29.22276868

**Authors:** Chrispin Chaguza, Anne M. Hahn, Mary E. Petrone, Shuntai Zhou, David Ferguson, Mallery I. Breban, Kien Pham, Mario A. Peña-Hernández, Christopher Castaldi, Verity Hill, Yale SARS-CoV-2 Genomic Surveillance Initiative, Wade Schulz, Ronald I. Swanstrom, Scott C. Roberts, Nathan D. Grubaugh

## Abstract

The chronic infection hypothesis for novel SARS-CoV-2 variant emergence is increasingly gaining credence following the appearance of Omicron. Here we investigate intrahost evolution and genetic diversity of lineage B.1.517 during a SARS-CoV-2 chronic infection lasting for 471 days (and still ongoing) with consistently recovered infectious virus and high viral loads. During the infection, we found an accelerated virus evolutionary rate translating to 35 nucleotide substitutions per year, approximately two-fold higher than the global SARS-CoV-2 evolutionary rate. This intrahost evolution led to the emergence and persistence of at least three genetically distinct genotypes suggesting the establishment of spatially structured viral populations continually reseeding different genotypes into the nasopharynx. Finally, using unique molecular indexes for accurate intrahost viral sequencing, we tracked the temporal dynamics of genetic diversity to identify advantageous mutations and highlight hallmark changes for chronic infection. Our findings demonstrate that untreated chronic infections accelerate SARS-CoV-2 evolution, ultimately providing opportunity for the emergence of genetically divergent and potentially highly transmissible variants as seen with Delta and Omicron.

## Introduction

Since the initial introduction of severe acute respiratory syndrome coronavirus 2 (SARS-CoV-2) in late 2019, subsequent coronavirus disease 2019 (COVID-19) waves have been predominantly driven by the emergence of variants with either enhanced transmissibility or the ability to evade human immune responses (Andrews et al., 2022; Dhar et al., 2021; Grubaugh and Cobey, 2021; Mlcochova et al., 2021; Takashita et al., 2022; Tegally et al., 2021; Volz et al., 2021). The SARS-CoV-2 lineage B.1.1.7, designated as Alpha by the World Health Organization (WHO), was the first named variant. Alpha was initially associated with a large cluster of cases in the United Kingdom before spreading globally (Volz et al., 2021). Analysis of the phylogenetic branch leading up to the B.1.1.7 clade revealed a faster evolutionary rate compared to the background evolutionary rate (Hill et al.); and the clade’s defining constellation of substitutions were associated with higher transmissibility compared to other lineages circulating at the time (Wu et al., 2020). Similar patterns of an unexpectedly long phylogenetic branch preceding a clade with increased transmissibility, disease severity or immune evasion have been observed multiple times with other variants: Beta (B.1.351), Gamma (P.1), Delta (B.1.617.2), and Omicron (B.1.529), causing extensive morbidity and mortality on national and international levels (Dhar et al., 2021; Naveca et al., 2021; Tegally et al., 2021; Viana et al., 2022).

Three mechanisms have been proposed for the emergence of genetically divergent SARS-CoV-2 variants: (1) prolonged human-human transmission in an unsampled population, (2) circulation in an unsampled zoonotic reservoir, and (3) chronic infection in an immunocompromised individual. Of these, chronic infection is the most plausible. Cryptic human-human transmission is unlikely to result in the increased evolutionary rate that is a hallmark of variants. Retrospective sequencing of cases may shorten the length of clade-defining branches, as was the case for Gamma (P.1), which likely emerged through stepwise diversification via multiple inter-host transmissions (Gräf et al., 2021). However, human-animal followed by animal-human transmission has been documented repeatedly, particularly in farmed mink populations (Oude Munnink et al., 2021), but there is no evidence to suggest that these events would produce monophyletic clades observed in most variants. Documented spillovers have not been associated with increased evolutionary rates nor have they led to community transmission. In contrast, a chronic SARS-CoV-2 infection in an immunocompromised individual is the best explanation for the emergence of Alpha based on evolutionary theory, when gaps in surveillance can be discounted (Hill et al.). Compared to between-host transmission, within-host dynamics can lead to increased evolutionary rates because the larger viral population is subject to fewer genetic bottlenecks (Braun et al., 2021; Lythgoe et al., 2021; Tay et al., 2022). This increases the selective impact imposed by a semi-functioning immune system relative to drift (Grenfell et al., 2004), and, in the case of SARS-CoV-2, increases the opportunity for recombination (Jackson et al., 2021). While extended community transmission associated with spillovers from animal reservoirs has not been observed, viruses from chronic infections have been detected in the broader community (Gonzalez-Reiche et al.; Wilkinson et al.). Despite this theoretical and epidemiological evidence that chronic infections could drive the emergence of variants, there is a dearth of detailed genomic analyses investigating the prolonged within-host evolutionary dynamics of the virus population in a chronically infected individual.

In this study, we investigate the intrahost genetic diversity and evolution of the SARS-CoV-2 B.1.517 lineage during 471 days of chronic infection of an immunocompromised individual suffering from advanced lymphocytic leukemia and B-cell lymphoma. Individuals who are immunocompromised are at an elevated risk of developing a persistent SARS-CoV-2 infection (**Table 1**) (Avanzato et al., 2020; Cele et al., 2022; Gandhi et al., 2022; Kemp et al., 2021; Maponga et al.; Weigang et al., 2021). An improved understanding of SARS-CoV-2 evolution during chronic infections could reveal targets for therapeutics to treat these infections and, as discussed above, curb the evolution and emergence of novel genetically divergent variants. Here, we characterize the longitudinal dynamics of viral RNA titers and infectious copies, intrahost genetic diversity, mutational spectrum and frequency, and recombination. We observe accelerated evolution of SARS-CoV-2 during the infection, marked by the emergence of distinct coexisting genotypes that could be designated as new lineages if transmitted to the community. We further demonstrate that the mutation accrual patterns of these genotypes resemble those seen in SARS-CoV-2 variants, including Omicron, and describe intrahost evolution dynamics to identify potential hallmark mutations associated with chronic infection. Together, our findings support the hypothesis that chronic infections could lead to the emergence of genetically divergent novel lineages with potentially high transmissibility and immune escape.

**Table 1:**
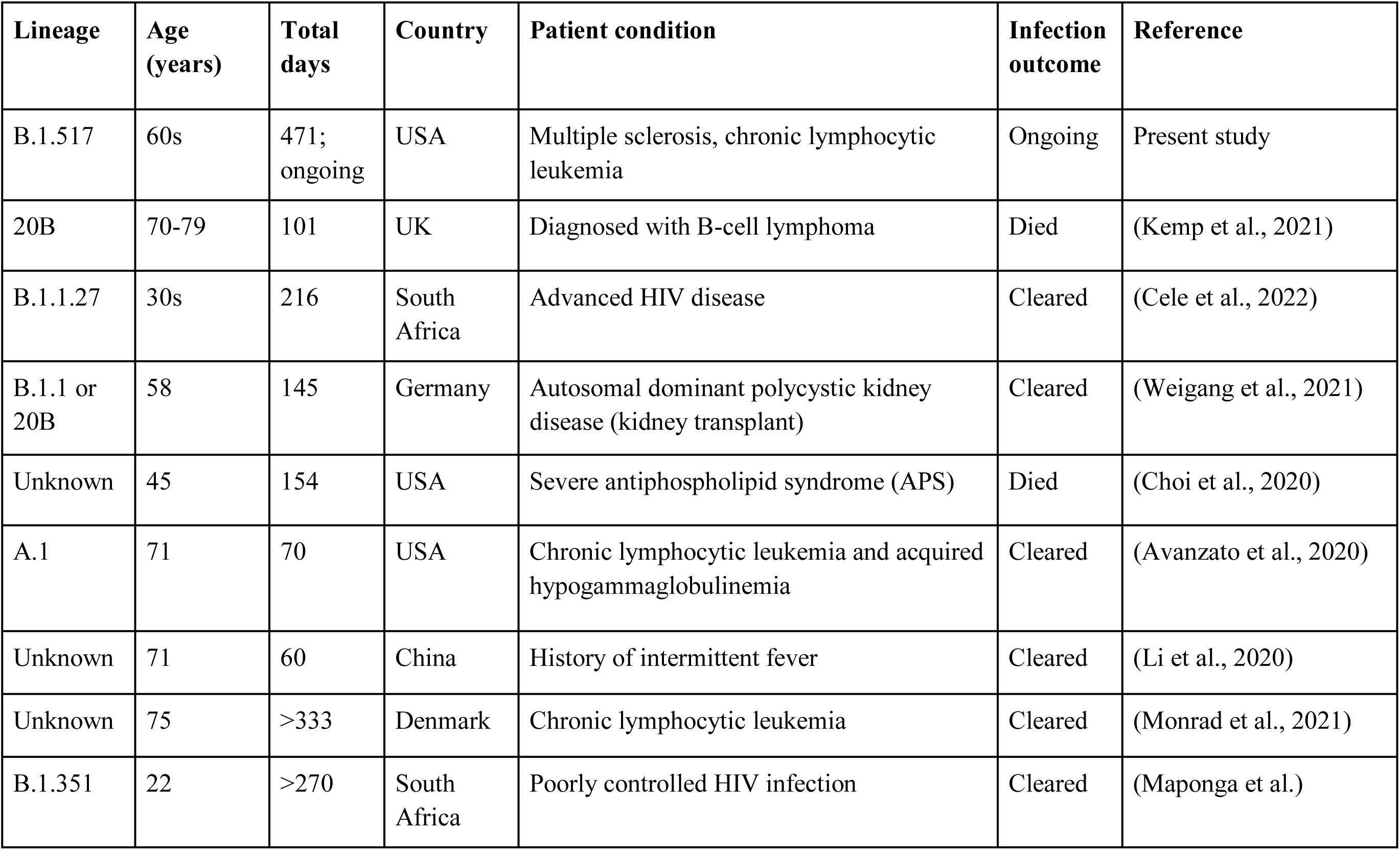
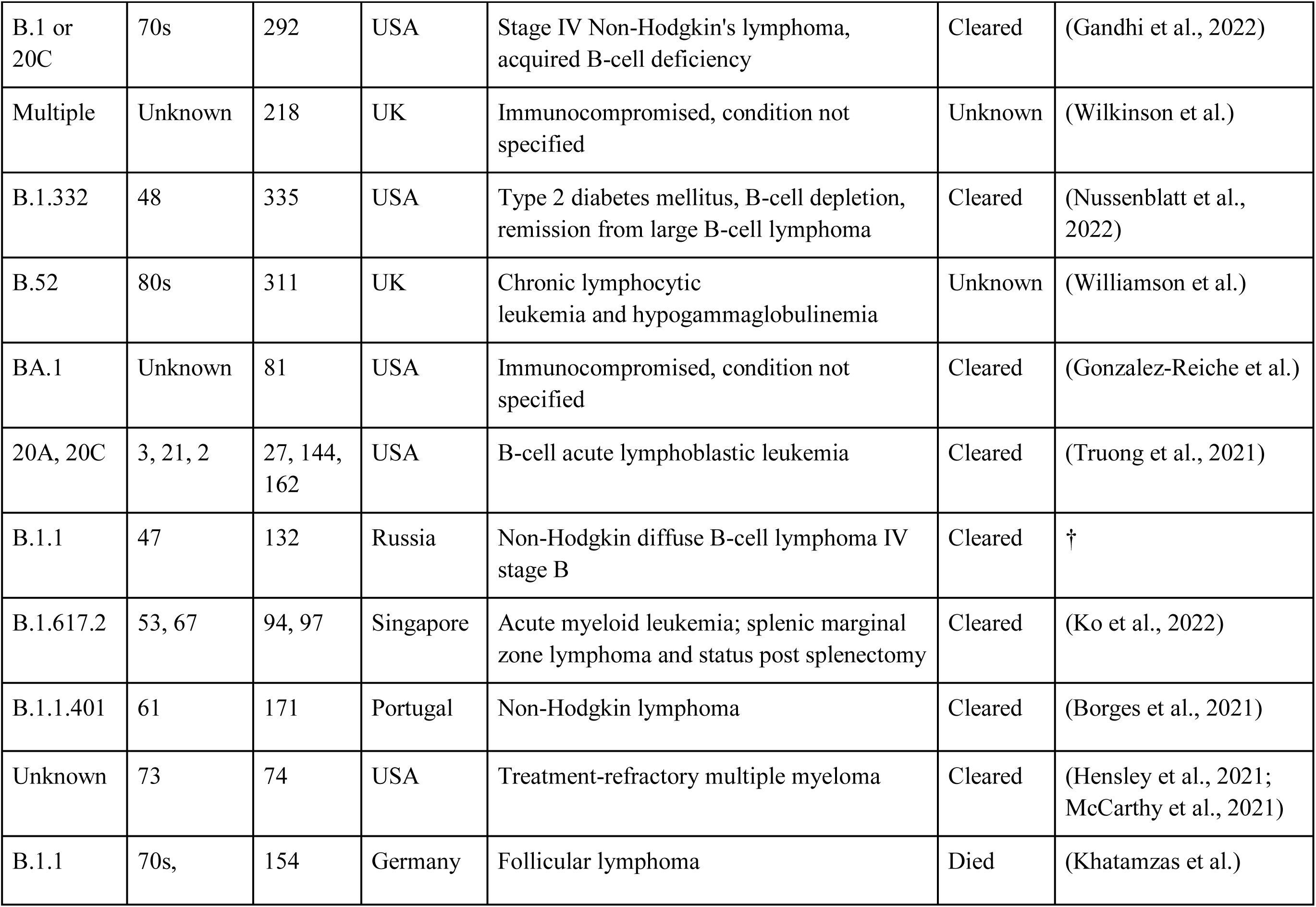
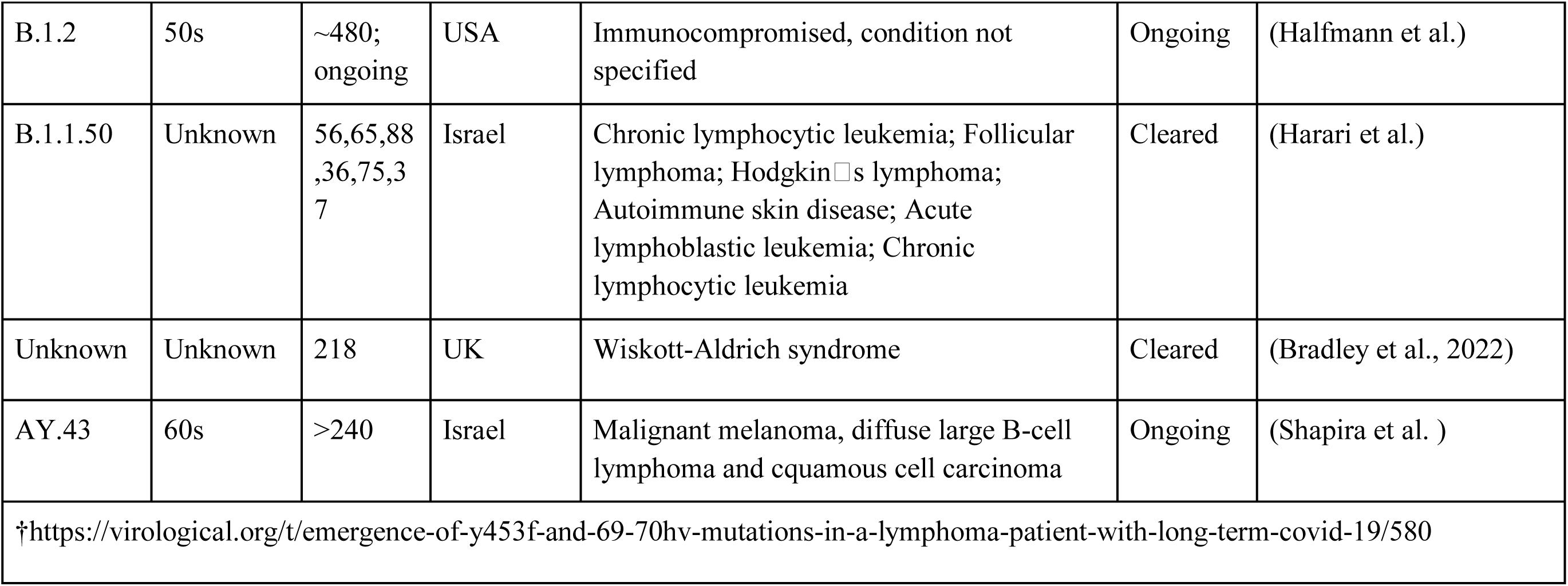
Summary of persistent SARS-CoV-2 infections in immunocompromised individuals reported to date.

## Results

### Chronic infection driving continued detection of B.1.517 in the United States

We identified the recurrent SARS-CoV-2 lineage B.1.517 in Connecticut, USA, extinct elsewhere in the US and globally, through our SARS-CoV-2 genomic surveillance initiative dataset (started in January 2021 with the emergence of Alpha) (**Figure 1A-B**). The B.1.517 lineage emerged in North America (likely in the US), in approximately early January 2020 (95% CI: November 2019 to March 2020) (**Figure 1C**). Following its emergence, B.1.517 spread to several US states and internationally, predominantly causing sporadic cases, except in Australia where an outbreak occurred (**Figure 1D**). Sequenced cases of B.1.517 in other countries remained sporadic and relatively low in frequency (**Figure 1A, B**). Lineage B.1.517 circulated until April 2021 in the US and other countries; however, we continued to detect B.1.517 in our genomic surveillance in Connecticut, USA until March 2022 (**Figure 1A, B**). We traced the recurrent B.1.517 sequences to an immunocompromised individual experiencing a chronic SARS-CoV-2 infection lasting 471 days at time of writing (**Figure 1E**, **Table 1**). Our surveillance system captured 30 nasal swabs from this individual and we sequenced SARS-CoV- 2 genomes from day 79 to 471 (February 2021 to March 2022).

**Figure 1:**
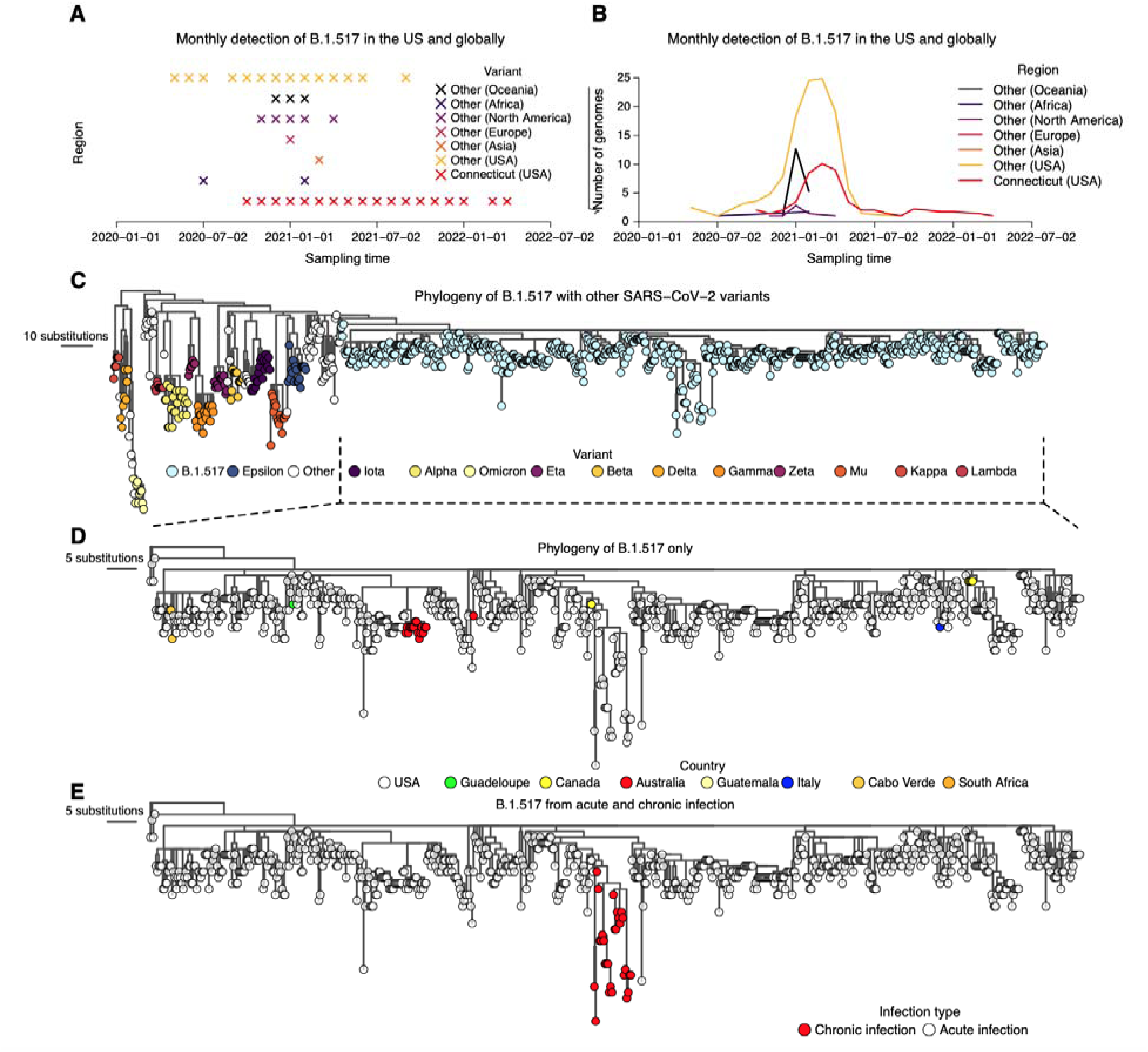
Genomic surveillance and phylogeny shows continued detection and genetic divergence of B.1.517 from chronic infection. (A) Monthly detection of B.1.517 (B.1.517 and B.1.517.1) variants in Connecticut (USA), other USA states, and elsewhere. **(B)** Total number of sequence genomes for the B.1.517 (B.1.517 and B.1.517.1) variants in Connecticut (USA), the rest of the USA, and elsewhere. **(C)** Maximum likelihood phylogeny of B.1.517 in the context of selected genomes from other variants. **(D)** Maximum likelihood phylogeny of all sequenced B.1.517 genomes showing country of origin. **(E)** Maximum likelihood phylogeny of all sequenced B.1.517 samples highlighting the genomes associated with the chronic infection and other contextual genomes from acute infection (although some could have been sampled from unknown chronic infections).

The patient found to be chronically infected with B.1.517 is in their 60s with a history of diffuse large B-cell lymphoma and underwent an allogeneic haploidentical stem cell transplantation in 2019. In early 2020, the patient’s disease relapsed and was started on a new chemotherapy regimen, ultimately requiring chimeric antigen receptor T-cell therapy in mid-2020. The patient was noted to have persistent but improving disease up until November 2020 when it started to relapse again. This is when the patient first tested positive for SARS-CoV-2 (November 2020, day 0), likely from a household contact that first tested positive for SARS-CoV-2 two days prior (**Figure 2A**). The patient was started on palliative radiation therapy on day 278 and was admitted three times from days 279 to 452 for malignancy-related complications. Clinical courses related to the infection are provided in **Figure 2A** and longitudinal immune parameters such as IgG serum levels as well as lymphocyte and T cell counts are provided in **Figure S1**. The patient’s IgG levels were within or near the reference range when receiving regular intravenous immunoglobulin therapy (IVIG) infusions until day 205, then the IgG levels dropped after IVIG treatment was suspended. The patient also had low lymphocyte, T cell, and IgA (non-detectable, data not shown in **Figure S1**) levels before and during the infection, consistent with their immunocompromised state.

**Figure 2:**
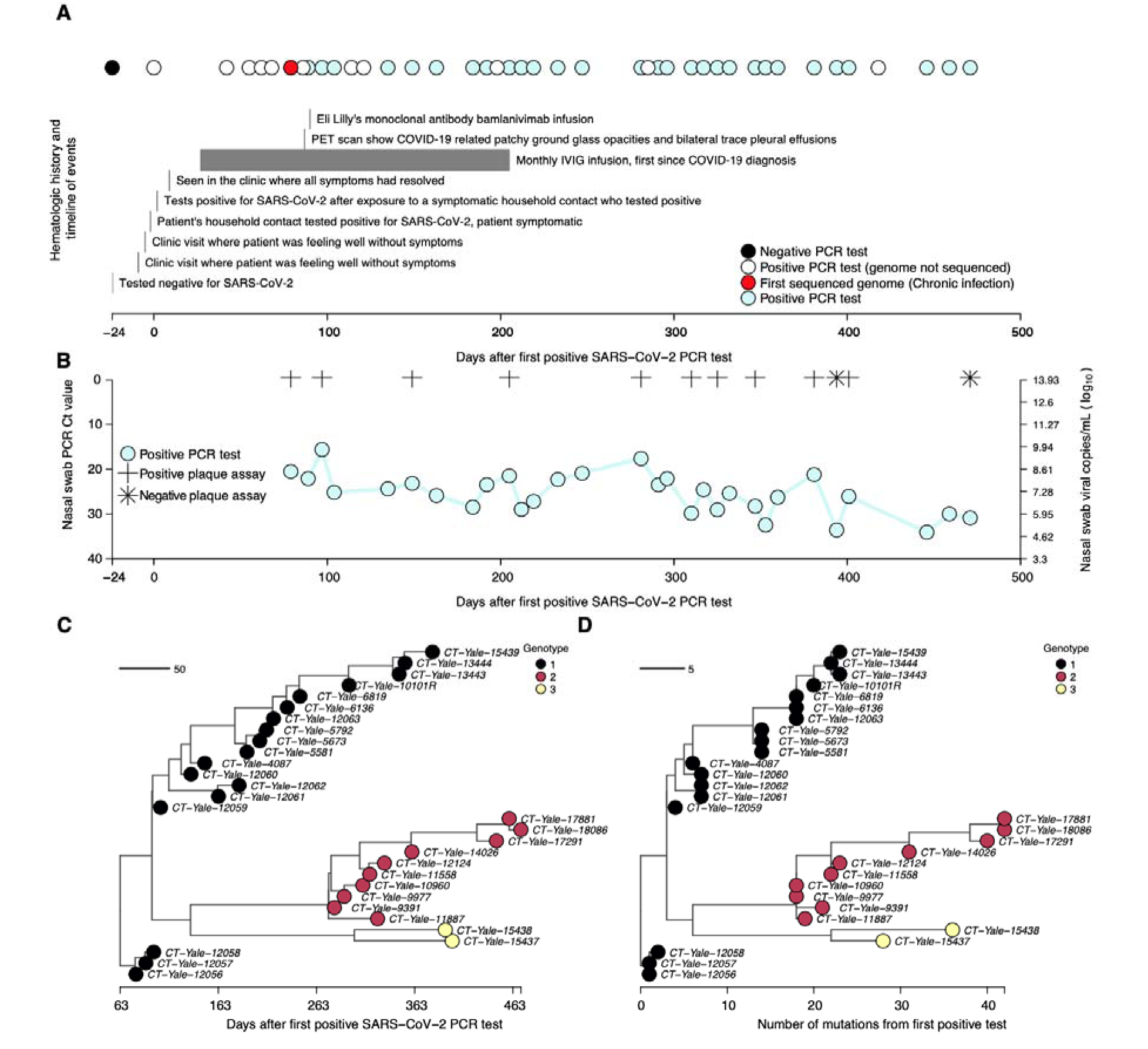
Molecular and virological assays showing isolation of infectious viruses with high copy numbers and the emergence and coexistence of distinct genotypes during the chronic infection. (A) Timeline showing clinical history of the patient from the earliest time they tested negative for SARS-CoV-2, the first positive test following household exposure by a symptomatic household contact who tested positive two days prior, until the last sampling point. Note that collection of samples was stopped due to the deteriorating condition of the patient, but the infection had not yet cleared. **(B)** Nasal swab RT-PCR cycle threshold (Ct) values for the samples available for whole genome sequencing showing high viral RNA copy numbers. Additionally, virus infectivity assays performed for selected samples revealed infectious virus at most sampling points. Additional information for the samples, including plaque assay results, are provided in **Table S1**. **(C)** Time-resolved phylogeny of the chronic infection samples with branch lengths scaled by the number of days since the first positive RT-PCR SARS-CoV-2 test. **(D)** Maximum-likelihood phylogeny of the chronic B.1.517 samples showing branch lengths scaled by the genetic divergence expressed as the number of accrued subsitutions over time. The phylogeny shows the intrahost emergence and persistence of multiple divergent genotypes.

Aside from the initial presentation of several days with mild upper respiratory tract symptoms not requiring oxygenation or hospitalization, the patient has remained asymptomatic for the duration of their SARS-CoV-2 infection. The only COVID-19 treatment the patient received was a bamlanivimab (LY-CoV555) monoclonal antibody infusion on day 90, after which the patient did not wish to obtain any additional COVID-19 therapies or vaccines. The patient continues to test positive for SARS-CoV-2 471 days and counting after the initial diagnosis.

### Persistently high viral RNA copies and infectious virus detected throughout the course of the chronic infection

To track the dynamics of the patient’s chronic infection, we quantified the viral RNA titers and investigated the virus infectivity from day 79 to 471 post diagnosis (February 2020 to March 2022; **Figure 2**, **Table S1**). The median number of days between successive samples was ∼14 days, 95% CI:8 to 20). We could not obtain samples from the patient prior to day 79 as they were collected before the establishment of our SARS-CoV-2 biorepository and genomic surveillance initiative. Though infection has not yet cleared at time of writing, sample collection was halted in March 2022 due to complications relating to the B-cell lymphoma disease, precluding further nasopharyngeal sampling.

We measured SARS-CoV-2 viral load using RT-PCR and performed whole-genome sequencing on 30 samples. We tested a subset of twelve for infectious virus and found the individual was infectious with high virus copies for almost the entire duration of their infection (**Figure 2B**).

Nasal swab samples collected from day 79 to 471 post diagnosis had a mean RT-PCR cycle threshold (Ct) of 25.50 (range: 15.6 to 34.1), equivalent to 3.10×10^8^ virus copies per mL (range: 7.30×10^4^ to 6.04×10^9^), though the copies numbers tended to decrease over time (**Figure 2B**, **Table S1**). Of the 12 swab samples that we tested for the presence of viable virus, infectious virus could be detected *in vitro* from ten sampling points (between days 79 and 401) but not on days 394 and 471, corresponding to samples with higher Ct values (33.6 and 30.9, respectively; **Figure 2B**, **Table S1**). However, the patient has been presumed to be asymptomatic for COVID- 19 after the resolution of the initial acute infection in November 2020, and all the patient’s admissions were secondary to malignancy. Given the sustained high viral load and infectiousness of viral particles in the nasopharynx, we concluded that the patient’s immune system was unable to suppress active SARS-CoV-2 replication throughout the infection **(Figure S1)**.

### Three distinct virus genotypes emerged during chronic infection

We hypothesized that SARS-CoV-2 from prolonged chronic infection would diversify into distinct populations, reflecting infection of spatially structured human cells and tissues. SARS-CoV-2 can infect diverse human cell populations and tissues (Delorey et al., 2021), similar to other pathogens including influenza virus (van Riel et al., 2006; Shinya et al., 2006) and bacterial pathogens (Jorth et al., 2015; Lieberman et al., 2016). To test this hypothesis, we constructed a phylogeny of the 30 longitudinally sequenced SARS-CoV-2 genomes from day 79 to 471 since the first positive SARS-CoV-2 test.

We identified three genetically divergent genotypes based on the phylogenetic clustering (numbered 1-3), which emerged and coexisted during the infection (**Figure 2C, D**). While we first sequenced genotype 1 on day 79, we cannot confirm that it was the founding genotype due to missing earlier samples. Genotype 1 accumulated up to 24 nucleotide substitutions (13 amino acid substitutions) through day 379 in a ladder-like evolutionary pattern. Genotype 2 diverged from genotype 1, with a maximum of 40 nucleotide substitutions (28 amino acid substitutions) from day 281 to 471. Genotype 3 also diverged from genotype 1 into two sister sub-genotypes sampled on day 394 to 401. The first sub-genotype accumulated 37 nucleotide substitutions (30 amino acid substitutions), while the second sub-genotype contained 29 nucleotide substitutions (27 amino acid substitutions) and diverged from each other on day ∼316 (95% CI: ∼288 to 336). These findings support our hypothesis that the founding B.1.517 virus independently diverged into coexisting genetically distinct populations.

Though the identified genotypes coexisted for the duration of the infection, the relative composition of the viral population changed over time (**Figure 2C, D**). We found that genotype 1 was dominant in nasal swabs from day 79 to 247; however, from day 281 to 471, the dominant genotype frequently switched between the three. From day 281 to 381, the sampled dominant genotype alternated between genotype 1 and 2 five times. Genotype 3 became dominant on days 394 and 401 before being replaced again by genotype 2 from day 446 to 471. The rapid and sometimes temporary replacement of genotypes during this infection suggested continual reseeding of the nasopharynx with distinct virus populations that likely independently evolved elsewhere in the body(Farjo et al.).

### SARS-CoV-2 evolution was accelerated during the chronic infection

The within-host evolutionary rate of microbes tends to exceed rates observed at the population level because of the absence of stringent bottlenecks imposed by transmission (Choi et al., 2020; Didelot et al., 2016). We thus hypothesized that the SARS-CoV-2 evolutionary rate during this chronic infection would be higher than the estimated global evolutionary rate. To test this hypothesis, we randomly sampled an equal number of genomes from the global dataset, ∼1 to 3 genomes per continent per month (n=2,539), for the WHO-designated SARS-CoV-2 variants and performed a regression of distance from the root of the phylogeny against time of sampling for the global dataset and the sequences from the chronic infection (**Figure 3**). We found that the evolutionary rate during the chronic infection was 35.55 (95% CI: 31.56 to 39.54) substitutions per year or ∼1.21×10^-3^ (95% CI: 1.07×10^-3^ to 1.34×10^-3^) nucleotide substitutions per site per year (s/s/y). This was ∼2 times higher than our estimated average global (all lineages) SARS-CoV-2 evolutionary rate (5.83×10^-04^ [95% CI: 5.56×10^-04^ to 6.11×10^-04^] s/s/y; **Figure 3A, B**; **Table S2**).

**Figure 3:**
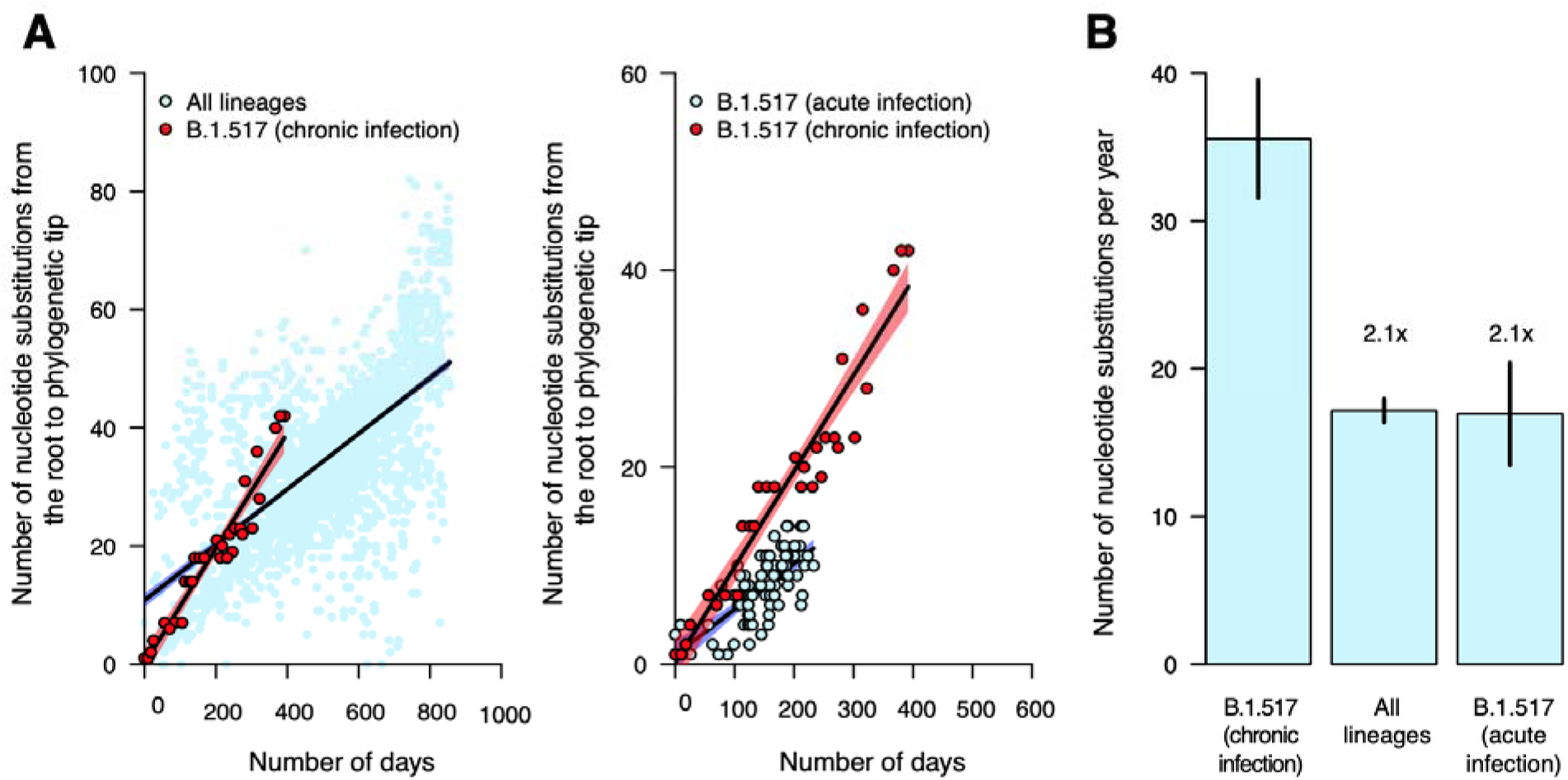
**Nucleotide substitution rates are faster during chronic infection than acute infection and the global evolutionary rate**. **(A)** Scatter plots showing relationship between phylogenetic root to tip distances, expressed as the number of nucleotide substitutions per site, and time as the number of days from the first sampled genome for the B.1.517 from chronic infection versus all SARS-CoV-2 lineages and other B.1.517 from acute infections. The data points associated with the chronic infection are coloured in red while those representing other variants are coloured in sky blue. The lines and shaded bands surrounding them represent the linear regression models fitted to the data points for the chronic infection data and other variants. **(B)** Bar graph showing the average mutation rates, expressed as the number of nucleotide substitutions per year for the chronic infection samples and other variants based on the regression coefficients (β) generated from the plots in panel A. Specific values for the evolutionary rates are shown in **Table S2**.

Our estimate for the global evolutionary rate, based on careful random sampling of representative genomes from GISAID per month per variant, is within the expected range of what is reported in other studies which use the same regression method (Fauver et al., 2020; Hill et al.). It is worth noting that estimates of the background rate of evolution vary due to different methodologies and downsampling used: 8×10^-4^ s/s/y is commonly used in phylodynamic analyses (Aggarwal et al.; Duchene et al., 2020; Nadeau et al., 2021), and the current (June 2022) Nextstrain estimate is approximately 9.9×10^-4^ s/s/y. However, even at these upper ends of the rate estimates, the rate of evolution in the chronic infection documented here is still faster. Our estimated evolutionary rate of this chronic B.1.517 infection is also ∼2 times higher than the evolutionary rate for the parental B.1.517 lineage (5.76×10^-04^ [95% CI: 4.58×10^-04^ to 6.94×10^-04^] s/s/y). These findings show that this chronic infection resulted in accelerated SARS-CoV-2 evolution and divergence, a mechanism potentially contributing to the emergence of genetically diverse SARS-CoV-2 variants, including Omicron, Delta, and Alpha.

### Increasing intrahost genetic diversity and variable gene-specific evolutionary rates during the chronic infection

Having detected three genotypes and observed the overall increased SARS-CoV-2 evolutionary rate during chronic infection, we hypothesized that intrahost virus genetic diversity would also increase over the course of infection. To test this hypothesis, we used deep sequencing to quantify the number of unique intrahost single nucleotide variants (iSNVs, i.e., “mutations”) present at >3% within sample frequency in each sample (**Figure 4A-D**). To validate the iSNV frequencies that we generated from whole genome amplicon-based sequencing, we sequenced the spike gene of a subset of the samples using unique molecular index (UMI)-tagged primers that improves the accuracy of iSNV detection (Zhou et al., 2015, 2021). We found a high concordance between the iSNV frequencies measured from our whole genome amplicon-based and UMI sequencing (median [β]: 0.999) (**Figure S2**, **Table S3**).

**Figure 4:**
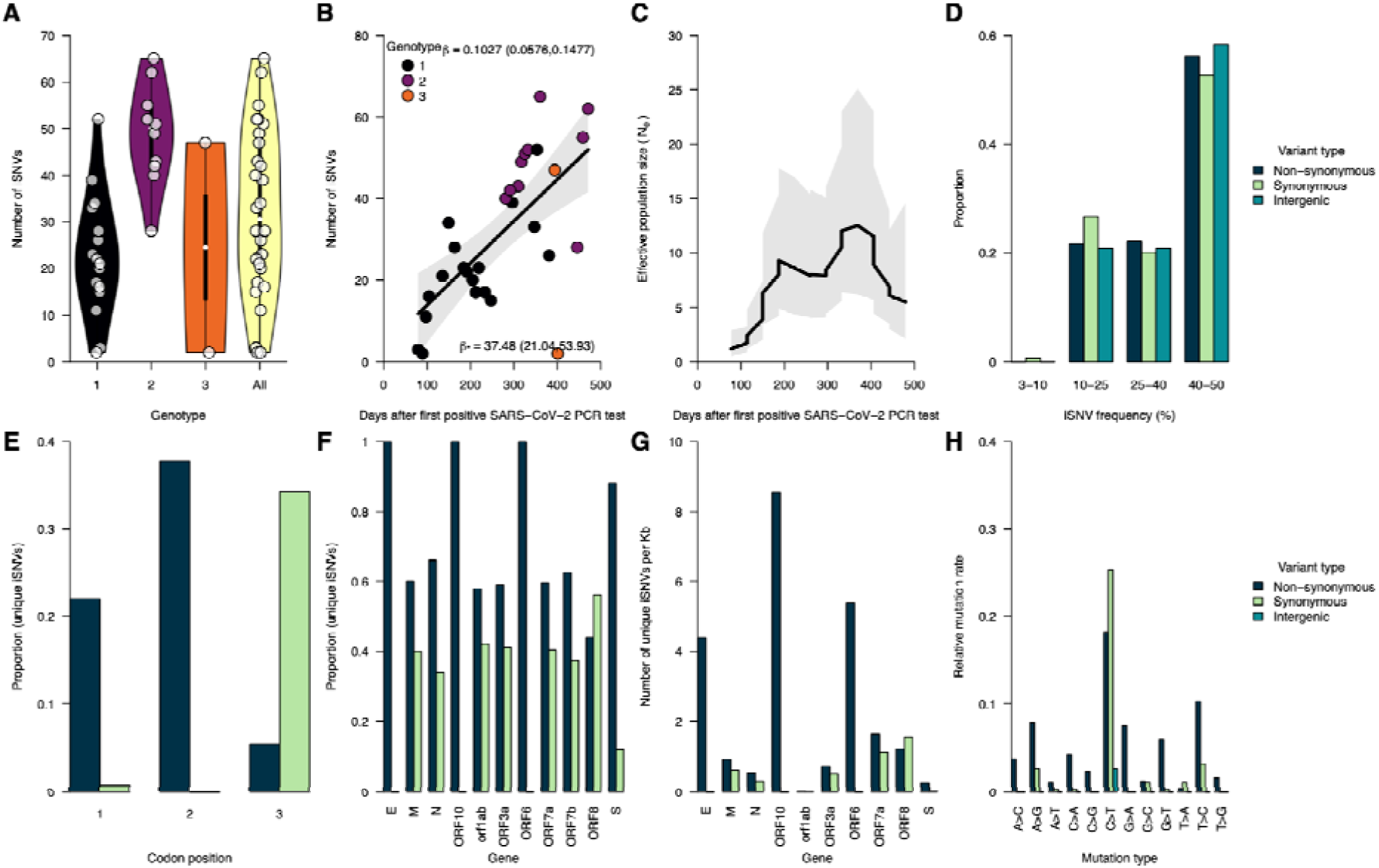
Increasing intrahost genetic diversity during chronic infection. (A) Number of intrahost single nucleotide variants (iSNVs) >3% frequency across all the samples and genotypes detected during the infection (see Figure 2C**, D**). **(B)** Number of iSNVs accumulated over time during the chronic infection. The black solid line represents a fitted linear regression. The regression coefficient (β), i.e., slope of the fitted line, represents the number accrued iSNVs per day while β* represents the number of iSNVs per year. **(C)** The effective population size (N_e_) pe day during the chronic infection, estimated based on the consensus genomes shown in Figure 2C**, D**. **(D)** Proportion of iSNVs binned at different frequencies, and stratified by variant or mutation type (intergenic, synonymous, and non-synonymous). **(E)** Number of unique iSNVs at different codon positions. **(F)** Proportion of unique iSNVs grouped by variant type to highlight potential selection across different SARS-CoV-2 genes. **(G)** Number of unique iSNVs per gene normalized by the gene length, to highlight variability in selection independent of gene size. **(H)** Mutation spectra showing the relative mutation rate across the SARS-CoV-2 genome stratified variant type. Additional information for all the identified mutations (intergenic, synonymous, and non-synonymous) are provided in **Data S1**.

The number of iSNVs increased over time across all three genotypes, and the viral effective population size (N_e_) fluctuated similarly. We observed a variable number of iSNVs per sample (mean: 32.07, range: 2 to 65). Genotype 2 comprised more iSNVs than genotype 1, which emerged earlier in the infection (**Figure 4A**). We used regression to assess the accrual rate of iSNVs and found a strong positive association between the number of iSNVs and sampling time (regression slope [β]: 0.013, 95% CI: 0.058 to 0.148 iSNVs per day) (**Figure 4B**). When scaled per year, we estimated an accrual rate of 37.48 (95% CI: 21.04 to 53.93) iSNVs per year, in agreement with the per-year evolutionary rate we estimated using the nucleotide substitutions detected in the consensus genomes (**Figure 3A, B**). Next, we measured changes in the N_e_ from the sequenced consensus genomes during the chronic infection using a coalescent Bayesian skyline model (Sagulenko et al., 2018). Our N_e_ estimates mirrored the dynamics of the number of unique iSNVs, especially in the early stages of the chronic infection, and peaked at ∼370 days post-diagnosis (**Figure 4C**). Finally, we characterized the iSNV frequencies and found that ∼55% of the iSNVs rose to frequencies of 40-50% during the infection (**Figure 4D**). These patterns were consistent for intergenic, synonymous, and non-synonymous iSNVs. Such high iSNV frequencies combined with the increasing number of iSNVs (**Figure 4B**, **Figure S3**) and N_e_ (**Figure 4C**) are in line with the coexistence of multiple genotypes within a sequenced sample and help to explain the consensus genotype switching that we described after day 281 of the infection (**Figure 2C, D**). Collectively, these data support our hypothesis that intrahost SARS-CoV-2 genetic diversity increased with time during the chronic infection to levels not typically reported during acute infections (Braun et al., 2021; Tonkin-Hill et al., 2021).

We investigated the potential impact of this diversity on virus evolution by analyzing the types of mutations and the gene-specific evolutionary rates during the chronic infection (**Figure 4E- H**). Stratifying the >3% iSNVs by codon position, we found that most occurred at the second and third codon positions (**Figure 4E**). Most of the substitutions at the first and second codon positions resulted in ∼22% and ∼38% non-synonymous changes, respectively, compared to 0.05% at the third codon. Because these changes could correspond to selection in different genes, we compared the proportion of synonymous and non-synonymous iSNVs. We hypothesized that the spike and other surface and membrane-associated proteins would have a higher abundance of non-synonymous amino acid changes than other genes as the principal targets of the host antibody-mediated immune response. Consistent with our hypothesis, we found a much higher abundance of non-synonymous changes than synonymous changes in the spike glycoprotein (abundance: ∼88%, *P=*5.03×10^-27^), envelope (abundance: ∼100%, *P=*0.013), and nucleocapsid (abundance: 66%, *P=*0.033) genes (**Figure 4F**). We also found a higher abundance of non-synonymous amino acid changes in nonstructural genes, namely ORF1ab polyprotein (abundance: ∼58%, *P=*0.001) and ORF10 (abundance: 100%, *P=*0.003). We normalized the estimates to account for the gene length to compare the abundance of synonymous and non-synonymous changes in different genes. Contrary to our hypothesis that the genes encoding the surface and membrane-associated proteins (spike, envelope, and membrane) would have the highest normalized frequency of non-synonymous changes, the highest frequencies occurred in the ORF10 gene, followed by ORF6 and envelope, while lower frequencies occurred in the other genes, including spike and membrane (**Figure 4G**, **Figure S4**). These differences suggested that other genes evolved faster than the spike gene during this chronic infection. Finally, the mutation spectra showed relatively higher C→T substitution rates, consistent with findings elsewhere (Popa et al., 2020; Tonkin-Hill et al., 2021), but we found that the C→T substitution equally resulted in synonymous and non-synonymous changes. In contrast, some substitutions, including A→G, G→A, G→T, and T→C, appeared to cause slightly more non-synonymous than synonymous changes (**Figure 4H**, **Figure S5**). Our findings suggest that the accelerated evolution during this infection resulted in variable accumulation of potentially advantageous substitutions across the SARS-CoV-2 genome.

### Persistently detected mutations associated with major variants

We hypothesized that specific iSNVs, particularly in the spike glycoprotein gene, were selectively advantageous and therefore were more prevalent than iSNVs in other genes. We tested this hypothesis by comparing the number of unique iSNVs across different samples between the spike and other genes (**Figure 4I**, **Figure S6**). Overall, we found no differences between the prevalence of unique spike and non-spike iSNVs across different samples (*P*=0.536). We then investigated if the frequency of the non-synonymous iSNVs across the samples was higher than intergenic and synonymous mutations. Again, we found similar prevalence of non-synonymous compared to intergenic (*P=*0.925) and synonymous iSNVs (*P=*0.704) and between intergenic and synonymous iSNVs (*P=*0.524). These findings demonstrated that the average persistence of iSNVs from different genes, regardless of their frequency of occurrence, were similar during the course of the infection.

While the distribution of mutations was not concentrated in the spike gene, some specific iSNVs could have been selectively advantageous and/or clinically important. Of the 114 iSNVs detected in more than one sample at >3% intrahost frequency, we found 23 changes in the spike gene of which ∼87% were non-synonymous (**Figure 5**, **Figure S7**). The three most common iSNVs, found in 22 of the 30 (88%) whole genome deep-sequenced samples, were (1) spike:Q493K, which may promote adaptation in murine infection models (Huang et al., 2021; Wilkinson et al.), (2) spike:T1027I, found in the Gamma variant, and (3) orf1ab:L2144P.

**Figure 5:**
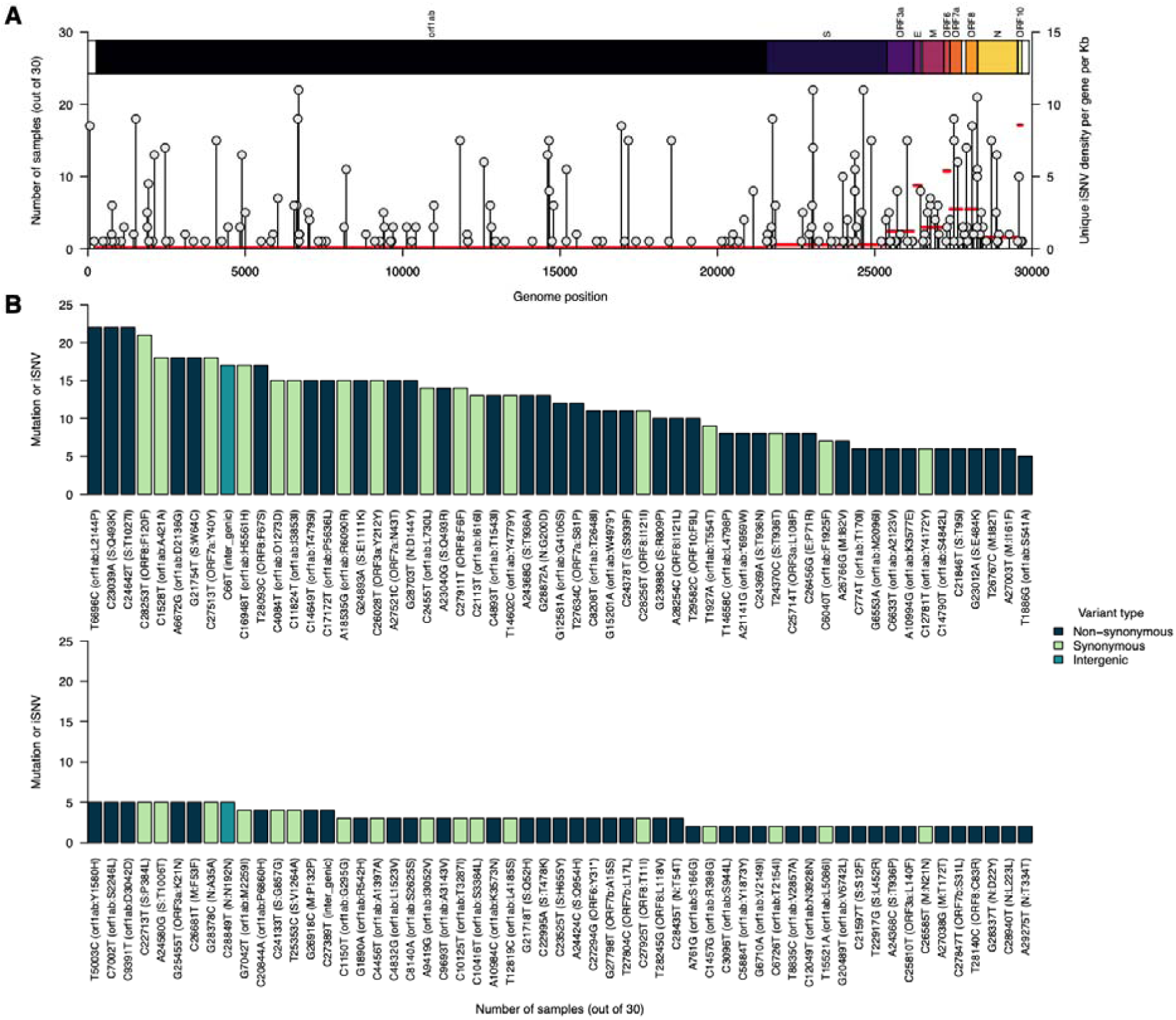
Several intrahost single nucleotide variants repeatedly detected during chronic infection. (A) Number of samples containing each unique intrahost single nucleotide variant (iSNV) and its position on the Wuhan-Hu-1 SARS-CoV-2 reference genome (GenBank: MN908937.3 or NC_045512.2). **(B)** The y-axis on the right side of the graph shows the number of iSNVs per gene and location of the gene in the reference genome. The y-axis labels represent iSNVs corresponding to specific nucleotide substitutions and position in the genome while the information within the blankets shows the specific amino acid changes, gene, and position in the gene. All the iSNVs are coloured by the variant or mutation type based on the Wuhan-1 SARS-CoV-2 genome sequence feature annotations (GenBank accession: MN908937.3). Additional information for all the identified mutations (intergenic, synonymous, and non-synonymous) are provided in **Data S1**.

We also detected several other iSNVs in the spike gene that have clinical relevance and/or are found in other variants. For example, the patient was treated with bamlanivimab (LY-CoV555) on day 90, and we detected three spike gene iSNVs associated with resistance to this antibody: Q493R, E484K, and L452R (Focosi et al., 2021; Jangra et al., 2021; Liu et al., 2022; Starr et al., 2021a; Weisblum et al., 2020). We detected spike:Q493R (found in Omicron) in 14 samples, with the first on day 97, one week after bamlanivimab treatment (**Figure 6**). The spike:E484K mutation (found in Beta, Gamma, Eta, Iota, and Mu) was detected in five samples from day 104 to 184. Finally, we detected spike:L452R (found in Delta, Epsilon, and Kappa) iSNVs in two samples, but not until day 401. These findings provide further evidence that clinically relevant mutations, such as those that confer resistance to antibodies and that are found in other variants, can evolve during the course of chronic infection.

**Figure 6:**
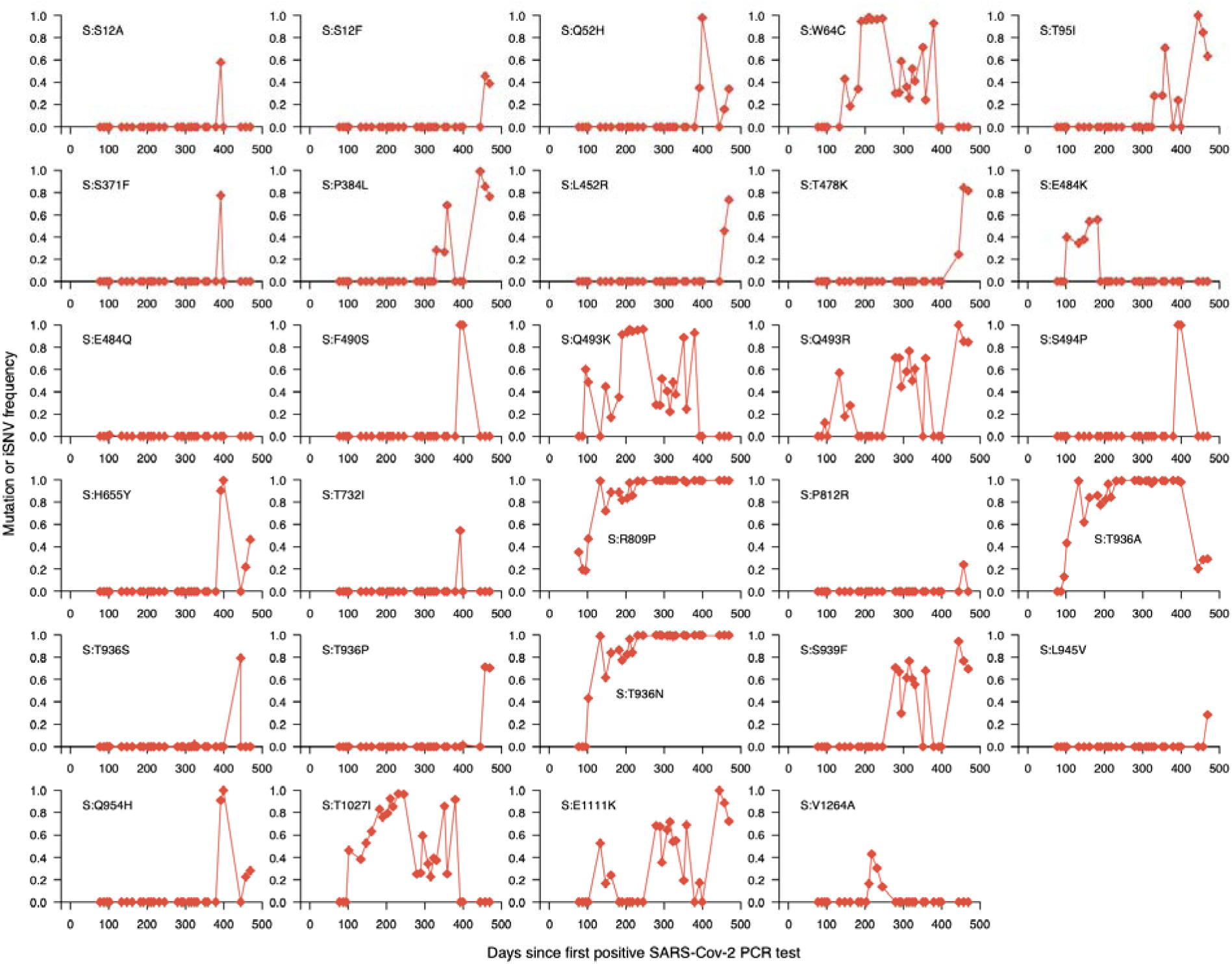
Fluctuating dynamics of intrahost single nucleotide variants in the spike gene during chronic infection. Temporal frequencies of twenty-nine non-synonymous intrahost single nucleotide variants (iSNVs) identified in the spike gene. Additional information for all the identified mutations (intergenic, synonymous, and non-synonymous) are provided in **Data S1**.

### Temporal mutational dynamics suggest unique hallmarks of chronic infection

To further understand spike gene iSNVs of potential significance during the chronic infection, we investigated temporal changes in their frequencies using deep sequencing validated with highly accurate UMI-based sequencing (**Figure 6**, **Figure S8**). We hypothesized that the frequency of beneficial non-synonymous spike gene iSNVs likely increased to reach near fixation during the infection. We found two iSNVs, spike:R809P between the fusion peptide and heptapeptide repeat sequence 1 (HR1) regions and spike:T936A/N in the HR1 region of the spike gene, which increased to near fixation throughout the infection, suggesting they were potentially beneficial to all the coexisting genotypes (**Figure 6**). Another notable spike mutation in the receptor-binding domain (RBD), spike:E484K, initially increased in frequency early in the chronic infection, as seen elsewhere (Choi et al., 2020), but was replaced by potentially fitter mutations and genotypes. Other spike iSNVs appeared to reach fixation, correlating with the detection of specific genotypes. These included, spike:1027I (genotype 1), spike:F490S (RBD; genotype 3), spike:Q52H (genotype 3), spike:P384L (RBD; genotype 2), and spike:493K (RBD; genotype 1). Outside of the spike gene, we detected other iSNVs that appeared to reach fixation: ORF1ab:T1543I (nsp3; genotype 2), ORF1ab:T2154I (nsp3; genotype 1 and 2), ORF1ab:S3384L (nsp5; genotype 3), ORF1ab:G4106S (nsp8; genotype 2), and ORF1ab:A3143V (nsp4; genotype 2; **Figure S8**), We conclude that most iSNVs fluctuate in frequency and rarely reach fixation. In contrast, a few spike iSNVs, which are novel and previously identified in variants and chronic infections elsewhere, attain fixation. We interpreted this as evidence for a selective advantage, possibly reflecting escape of the host antibody-mediated immune response, but we could not rule out other neutral evolutionary processes.

### No evidence for intrahost recombination during chronic infection

The long duration of this infection, which spanned the emergence of multiple variants (e.g., Alpha, Delta, Omicron), provided favorable conditions for recombination. The occurrence of recombination in the SARS-CoV-2 genome has been demonstrated (Jackson et al., 2021; Wertheim et al.). Therefore, we hypothesized that recombination may have occurred during the chronic infection between coexisting B.1.517 genotypes and between B.1.517 genotypes and other circulating variants transiently causing undetected co-infections. To test this hypothesis, we conducted a recombination analysis of the consensus genomes generated from the persistent infection samples. Since multiple genotypes emerged during the chronic B.1.517 infection, we first investigated the occurrence of intrahost recombination amongst these genotypes during the infection. We then tested whether recombination occurred between the B.1.517 chronic infection strains and other non-B.1.517 variants detected in Connecticut, USA, especially Delta and Omicron lineages. We found no evidence of recombination between the chronic B.1.517 genotypes or other variants. These findings suggested that the emergence of multiple genotypes during the B.1.517 infection evolved independently from the ancestral B.1.517 following infection due to random mutational processes rather than intrahost recombination.

## Discussion

In our comprehensive genomic investigation, we characterized the intrahost genetic diversity and evolution of SARS-CoV-2 during a chronic infection that has persisted for over a year. Our phylogenetic analysis, based on sequencing 30 nasal swab samples from days 79 to 471 post diagnosis, revealed accelerated SARS-CoV-2 evolution and the emergence and coexistence of multiple genetically distinct genotypes – a finding not reported in other studies reflecting the duration of the infection and longitudinal sampling. These distinct genotypes appeared to emerge as early as within the first three months of the infection, although new genotypes were detected after nearly ten months, suggesting multiple novel variants may simultaneously emerge and potentially spread from the same immunocompromised individual over a longer sampling period. Supporting this point, we detected high viral RNA copies and infectious virus throughout the duration of infection, even though the patient remained asymptomatic for COVID-19. A strength of this study was our ability to collect samples for a substantial portion of the infection because it enabled us to document the patient’s prolonged infectiousness. This critical finding could potentially be missed if data from chronic infections collected over shorter time scales were used. Our study provides evidence that chronic SARS-CoV-2 infections could be a source for the emergence of genetically diverse variants capable of causing future COVID-19 outbreaks.

During this infection, the viral population accrued twice as many nucleotide substitutions per year as those driving acute infections. Our findings support the prevailing hypotheses that chronic infections in immunocompromised individuals could be the most likely mechanism driving the unpredictable emergence of genetically diverse SARS-CoV-2 variants (Callaway, 2022; Kupferschmidt, 2021; Lemieux and Luban, 2022; Mallapaty, 2022). We have shown that the accelerated evolution observed in other SARS-CoV-2 variants such as Omicron and Alpha, which are considered to have emerged during unknown chronic infection, are consistent with the accrual of nucleotide substitutions demonstrated in our study (Cele et al., 2022; Hill et al.; Viana et al., 2022). Although previous studies have reported that most SARS-CoV-2 populations associated with chronic infections are homogenous, we found multiple genotypes coexisting throughout a single infection. The prolonged infectiousness of this patient demonstrated that a single chronic infection could cause onward transmission of multiple genetically distinct SARS-CoV-2 variants into the broader population. This could be especially problematic as many chronic infections, as was the case with this patient, remain mostly asymptomatic for COVID-19 and may feel well enough to resume regular interactions with other people. The direct, onward transmission of B.1.616 and BA.1 lineages from chronic infections has already been documented (Gonzalez-Reiche et al.; Wilkinson et al.). Therefore, it is possible that the simultaneous emergence of divergent Omicron sub-lineages (e.g. BA.1 and BA.2) could have been from a single long chronic infection (Cele et al., 2022; Viana et al., 2022). Altogether, our findings suggest that a novel variant could evolve into genetically divergent forms during a single chronic infection.

We speculate that the emergence and disappearance of multiple genotypes reflect virus competition in the nasopharyngeal niche and/or isolated evolution in different compartments of the respiratory tract or other tissues. These compartments may act as reservoirs for the genotypes and reseed them into the nasopharynx, leading to their fluctuating dynamics that can be observed in the swab material. A similar phenomenon has been reported in studies of acute SARS-CoV-2 infection (Farjo et al.) and chronic bacterial infections (Jorth et al., 2015; Lieberman et al., 2016; Viberg et al., 2017). Infection of multiple tissues leads to spatial isolation and niche partitioning, which ultimately reduces intrahost competition between distinct genotypes and promotes the coexistence of numerous genotypes over longer timescales (Jorth et al., 2015; Lieberman et al., 2016). Niche partitioning is plausible because different SARS-CoV-2 variants preferentially infect different cell types (Meng et al., 2022). Recent studies have demonstrated that Omicron has evolved a shift in the cellular tropism towards cells expressing transmembrane protease serine 2 (TMPRSS2), allowing it to more effectively infect upper airway cells as compared to endothelial cells of the lung, unlike other lineages (Meng et al., 2022). This process may similarly occur during accelerated SARS-CoV-2 evolution in chronically infected persons. While intrahost recombination may accelerate intrahost divergence (Jackson et al., 2021; Wertheim et al.), we did not find evidence for recombination leading to the distinct genotypes found during this chronic infection. This might be an indication for the separated spatial distribution of the viral populations as recombination events would be expected if different genotypes were to be found in the same tissues and cells. The differences in transmission fitness and cellular tropism among these genotypes requires further investigation.

The SARS-CoV-2 spike is a homotrimeric transmembrane glycoprotein critical for receptor recognition, cell attachment and entry, and an immunodominant target for host immune responses (Harvey et al., 2021). We found a higher abundance of non-synonymous than synonymous changes in five of the eleven SARS-CoV-2 genes, including the spike. This suggests positive selection during the course of the infection. Interestingly, although we detected the spike:E484K substitution, it did not reach fixation and lasted for approximately three months following bamlanivimab (LY-CoV555) treatment. This suggests that despite E484K being associated with antibody evasion (Greaney et al., 2021; Jangra et al., 2021; Starr et al., 2021b), it is not necessarily a hallmark of chronic infection involving an immunocompromised person, consistent with previous reports (Wilkinson et al.). However, we propose that iSNVs that reached near fixation (spike R809P and T936A/N) could be selectively advantageous during chronic infection. Furthermore, we hypothesize that spike Q493K/R mutation could be important for chronic SARS-CoV-2 infections (Choi et al., 2020; Peacock et al., 2021; Wilkinson et al.), even though neither became fixed in our study because they were on different genotypes. By validating the iSNV frequencies using a UMI-based sequencing approach (Primer ID) which helps to remove PCR artifacts (Zhou et al., 2015, 2021), our findings provide a robust assessment of intrahost evolutionary dynamics during chronic infection.

Chronic SARS-CoV-2 infections have been reported in individuals with compromised immunity due to a myriad of factors, including advanced HIV, cancer, organ transplant recipients, kidney disease, and autoimmune disorders (Avanzato et al., 2020; Cele et al., 2022; Choi et al., 2020; Gandhi et al., 2022; Kemp et al., 2021; Maponga et al.; Nussenblatt et al., 2022; Weigang et al., 2021). These infections may drive the rapid evolution of SARS-CoV-2 variants, including from lineages considered to be less virulent, which may spread into the broader population after acquiring mutations promoting increased intrinsic transmissibility and immune escape. As seen with Alpha, which cryptically evolved for >1 year before causing a global epidemic (Viana et al., 2022), variants that are likely to cause major future outbreaks could be “lying in wait” in unknown chronic infections. Therefore, control measures for COVID-19 should not only include decreasing cases associated with prevailing variants but also identifying and treating chronic infections to disrupt the potential emergence of novel variants. Moreover, since immunocompromised individuals typically exhibit greater healthcare-seeking behavior, implementation of proactive surveillance of chronic SARS-CoV-2 infections could substantially limit the rate of SARS-CoV-2 evolution (Collie et al., 2022; Dennehy et al., 2022). Considering that novel variants can emerge and transmit globally from anywhere, as seen with Omicron (Viana et al., 2022), these measures need global adoption to maximize their benefits.

In this study, we have shown accelerated intrahost evolution and genetic diversity of SARS-CoV-2 during a chronic infection lasting more than one year. Our findings show evolutionary patterns resembling those seen leading up to the Alpha and Omicron variants, highlighting the critical role of chronic SARS-CoV-2 infections in the emergence of novel variants. Therefore, we recommend proactive genomic surveillance of immunocompromised individuals to identify and treat potential chronic infections early, increased global equitable access and uptake of primary and booster COVID-19 vaccine regimens, and continued investment in the development of pan-β-coronavirus vaccines (Burton and Topol, 2021; Morens et al., 2022), to reduce the likelihood of chronic infections (Dennehy et al., 2022). These strategies could halt the accelerated evolution of SARS-CoV-2 seen in chronically infected individuals, disrupting the emergence of genetically divergent and more transmissible variants, ultimately averting mortality, morbidity, and the tremendous economic impacts of strict COVID-19 prevention and control measures.

### Limitations of the study

Although we have performed a detailed genomic investigation of the intrahost evolution and genetic diversity during the chronic infection, a potential limitation of our study is that we have characterized a single case. However, we have utilized other published case studies of chronic SARS-CoV-2 infection to contextualize our findings and understand commonalities and differences between infections. Additionally, we did not compare the antibody neutralization susceptibility of different intrahost genotypes emerging during the chronic infection. Therefore, future studies of chronic infections, especially those utilizing prospectively collected samples, should include longitudinal and parallel samples to monitor several immune parameters such as antibody levels and immune cell composition as well as serum samples for neutralization assays to generate additional insights on the persistence and evolution of multiple genetically distinct genotypes in the same host. For this study, we did not have access to this additional information including Human leukocyte antigen (HLA) haplotype data which would have been valuable in evaluating the contribution of the host’s immune system to the emergence of the observed genetic diversity of the viral population.

## Methods

### Ethics statement

This study was approved by the Yale University Human Research Protection Program Institutional Review Board (IRB Protocol ID: 2000031415). Informed consent was obtained from the participant to take part in the study and to have the results of this work published. The coded numbers presented in the tables and figures are not identifiable to the patient.

### PCR testing and whole-genome sequencing

Nasal swabs collected from the anterior nares or nasopharynx of confirmed SARS-CoV-2 positive individuals were routinely tested by the Yale New Haven Hospital COVID-19 and Clinical Virology Laboratories. We received remnant samples that were used for diagnostic testing. We used the MagMAX viral/pathogen nucleic acid isolation kit to extract nucleic acid from 300 μl of the collected sample by eluting in 75 μl of the elution buffer. We then extracted nucleic acid and tested it for SARS-CoV-2 RNA using a “research use only” (RUO) RT-qPCR assay using the CDC nucleocapsid gene target (N1) primer and probe set (Vogels et al., 2020). We converted the resulting N1 RT-PCR Ct values into SARS-CoV-2 RNA copies using a standard curve (Vogels et al., 2021).

We used the Illumina COVIDSeq Test RUO version to sequence samples with N1 PCT Ct values ≤35. We used ARTIC V3, V4, and V4.1 primer schemes for amplicon generation (https://github.com/artic-network/artic-ncov2019/tree/master/primer_schemes/nCoV-2019). We used a slightly modified sequencing protocol involving lowering the annealing temperature to 63°C when generating the amplicons and shortening the tagmentation step to 3 minutes. We pooled and cleaned the final libraries before DNA quantification using the Qubit High Sensitivity dsDNA kit (Life Technologies). The generated libraries were deep-sequenced using 2×150 bp paired-end reads on an Illumina NovaSeq at the Yale Center for Genome Analysis. At least one million paired-end reads were generated for each sample. The sequencing data were demultiplexed and processed, including converting base call (BCL) to FASTQ formats and trimmer adapter sequences, using Illumina bcl2fastq pipeline (v2.20.0). To generate consensus SARS-CoV-2 whole genomes, we aligned the reads to the Wuhan-Hu-1 reference genomes (GenBank: MN908937.3 or NC_045512.2) using BWA-MEM (version 0.7.15) (Li, 2013) to generate indexed and sorted binary alignment map (BAM) files. We trimmed adaptors, masked primers and generated consensus base calls for the BAM files based on simple majority >60% base frequency using iVar (version 1.3.1) (Grubaugh et al., 2019) and SAMtools (version 1.7) (Li et al., 2009). We defined ambiguous base calls as nucleotide sites containing <20 unique mapped reads. To validate the sequencing runs, we sequenced negative controls, and in all cases consisted of >99% sites with Ns. We selected sequences containing >70% of non-N base calls for submission to GISAID. We assigned the SARS-CoV-2 lineages using Pangolin (version 3.1.17) (O’Toole et al., 2021; Rambaut et al., 2020).

### Testing for infectious virus in nasopharyngeal swabs samples

To determine if the samples that test positive for viral RNA also contain infectious virus, we tested whether cell lines can be infected through nasopharyngeal swab material. For this, we chose twelve samples (40%) collected throughout the course of infection and available from the biorepository. For this, transmembrane protease serine 2 (TMPRSS2)-ACE2-VeroE6 kidney epithelial cells were cultured in Dulbecco’s Modified Eagle’s Medium (DMEM) supplemented with 1% sodium pyruvate (NEAA) and 10% Fetal bovine serum (FBS) at 37 °C and 5% CO_2_.

The cell line was obtained from the American Type Culture Collection (ATCC) and tested negative for Mycoplasma contamination. Briefly, 250 µl of serial fold dilutions of sample material obtained from nasopharyngeal swabs in viral transport medium were used to infect TMPRSS2-ACE2-Vero E6 cells for 1 hour at 37 °C for adsorption. We overlaid the cells with Minimum Essential Medium (MEM) supplemented with NaHCO_3_, 4% Fetal Bovine Serum (FBS) and 0.6% Avicel RC-581. We resolved the plaques at 72 h post-infection by fixing them in 10% formaldehyde for 30 minutes, followed by 0.5% crystal violet in 20% ethanol staining. We then rinsed the plates in water, and assessed presence or absence of plaques. All experiments were carried out in a biosafety level 3 and biocontainment (BSL3) laboratory with approval from the Yale Environmental Health and Safety (EHS) office.

### Phylogenetic reconstruction and recombination analysis

For the phylogenetic analysis, we masked the sites in the 5′ (position 1 to 265) and 3′ (position 29,675 to 29,903) genomic regions, which are typically poorly sequenced and are known to bias the phylogeny. To understand the genetic relationship of the consensus SARS-CoV-2 genomes from the chronic infection and other WHO designated SARS-CoV-2 variants (https://www.who.int/activities/tracking-SARS-CoV-2-variants), we constructed phylogenetic trees with branches resolved by time and genetic divergence, i.e., number of mutations, using the Nextstrain pipeline (version 3.0.3) (Hadfield et al., 2018). We used Nextalign (version 1.10.2) (https://github.com/neherlab/nextalign) and Augur (version 11.1.2 )(Huddleston et al., 2021), implemented in the Nextstrain pipeline, to filter out the genomes based on sampling dates, construct maximum likelihood phylogenies using IQ-TREE (version 2.0.3) (Nguyen et al., 2015), refine and reconstruct mutations on the phylogeny, and estimate the effective population size (N_e_). The last was based on the Coalescent Bayesian Skyline model using Treetime (version 0.8.1) (Sagulenko et al., 2018). Finally, interactive visualization was undertaken using Auspice (version 2.23.0) (https://auspice.us/) (Hadfield et al., 2018).

For other variants, we randomly selected up to three contextual SARS-CoV-2 genomes per month per lineage (Pangolin) from the GISAID database (Shu and McCauley, 2017) using dplyr (https://github.com/tidyverse/dplyr), and phylogenies generated using the same approach. We processed and visualized phylogenetic trees, including calculating root-to-tip distances, using ape (version 5.6.2) (Paradis et al., 2004) and phytools (version 0.7.70) (Revell, 2012). We generated plots showing the location of mutations in the nucleotide sequence alignment using snipit (https://github.com/aineniamh/snipit).

To test for potential recombination, we used 3SEQ (version 1.7) (Lam et al., 2018) to check for potential recombination, first amongst the genomes from the chronic infection and also in comparison with randomly selected genomes belonging to other SARS-CoV-2 variants detected in Connecticut, USA, over the course of the chronic infection.

All the statistical analyses, including regression, were done using R (version 4.0.3) (R Core Team, https://www.R-project.org/).

### Intrahost evolution and genetic diversity analysis

To investigate the intrahost evolution and genetic diversity during chronic infection, we first used ’MarkDuplicates’ in Picard (version 2.18.7) to identify duplicate reads in the BAM files of each sample (http://broadinstitute.github.io/picard/). We calculated the per-base sequencing depth using genomecov option in BedTools (version 2.30.0) (Quinlan and Hall, 2010). The bcftools (version 1.11-99-g5105724) (Danecek et al., 2011; Li, 2011) were used to generate variant calls for each sample using the reconstructed ancestral sequences for the chronic infection samples using the ’ancestral’ option in the Augur pipeline (version 11.1.2) (Huddleston et al., 2021), which uses Treetime (version 0.8.1) (Sagulenko et al., 2018). We specified a maximum depth of 1,000,000 with a minimum of 50 mapped reads per nucleotide site to infer variant calls. We used bcftools to calculate iSNV frequencies per sample and merge variant call files for different samples for annotation with vcf-annotator (version 0.7) (https://github.com/rpetit3/vcf-annotator) using the reconstructed ancestral sequence generated from the phylogeny of the chronic infection genomes and annotated using the Wuhan-1 SARS-CoV-2 reference genome. We compared the commonness of iSNVs across different samples based on the variant type (intergenic, synonymous, and non-synonymous) and gene using the unpaired two-sample Wilcoxon test or Wilcoxon rank sum test. We analyzed and visualized the presence and absence of mutations and their dynamics using R (version 4.0.3).

### Primer ID sequencing using unique molecular identifiers

UMI-guided deep sequencing was done using the previously published Primer ID next-generation sequencing protocol to sequence the SARS-CoV-2 viral genomes extracted from the specimens (Zhou et al., 2015, 2021). We used two sets of multiplexed UMI-tagged primers targeting SARS-CoV-2 ORF1ab (nsp12) and the spike gene. The cDNA and first-round PCR primers are provided in **Table S3**. After two rounds of PCR amplification, purified and pooled libraries were deep-sequenced using MiSeq 300 base paired-end sequencing. Sequencing data were first processed using the Illumina bcl2fastq pipeline to convert BCL to FASTQ and trimmer adapters (v2.20.0), followed by the TCS pipeline (v2.5.0) (https://www.primer-id.org/tcs) to de- multiplex for sequencing regions, construct template consensus sequences (TCS). We used BWA-MEM (version 0.7.15) (Li, 2013) to map the TCSs against the reconstructed ancestral

B.1.517 sequence for the chronic infection generated from the phylogeny of the chronic infection genomes and annotated using the Wuhan-1 SARS-CoV-2 reference genome (GenBank: MN908937.3), bcftools (version 1.11-99-g5105724) (Danecek et al., 2011; Li, 2011) to generate variant calls, calculate iSNV frequency and merge the variant files, and vcf-annotator (version 0.7) (https://github.com/rpetit3/vcf-annotator) to annotate the merged variants.

### Clinical Data

Information on clinical history and treatment was obtained from Yale New Haven Hospital. Longitudinal measurements of immune parameters (IgG levels, lymphocyte and T cell counts) were taken from chart review and obtained by standard clinical operation procedures.

### Data and code availability

A summary of the SARS-CoV-2 samples is available in Table S1. All other data and code are available at https://github.com/grubaughlab/2022_paper_chronic_infection.

## Acknowledgements

We would like to thank the Yale New Haven Health COVID-19 testing enterprise for collecting and testing samples and the health care workers for supporting the patients, and the patients for contributing samples. We also thank J.T. McCrone, P. Jack, and S. Taylor for technical discussions about the methodologies. This work was supported by the Centers for Disease Control and Prevention (CDC) Broad Agency Announcement #75D30120C09570 (N.D.G.). This work was also supported by NIH award R01-AI140970 to R.S. This research received infrastructure support from the UNC CFAR (P30-AI050410), and the UNC Lineberger Comprehensive Cancer Center (P30-CA016086). We acknowledge the support of the UNC High Throughput Sequencing Facility.

## Author contributions

CC, MP, RCS, and NDG conceived the study; RCS, DF, WS, and NDG collected the clinical data and/or samples; MIB, AMH, and NDG performed DNA extraction and sequencing library preparation; CC, AMH, MP, KP, CCa, and NDG performed the whole genome sequencing and analysis; SZ and RIS performed Primer ID sequencing; AMH and MAPH performed plaque assay experiments; CC, MP, AMH, and NDG designed the analysis methods and analyzed the data; SCR wrote the IRB protocol; CC, MP, AMH, RCS, and NDG drafted the manuscript; NDG secured funds and supervised the project; All authors reviewed and approved the manuscript.

## Supplementary information

**Figure S1:**
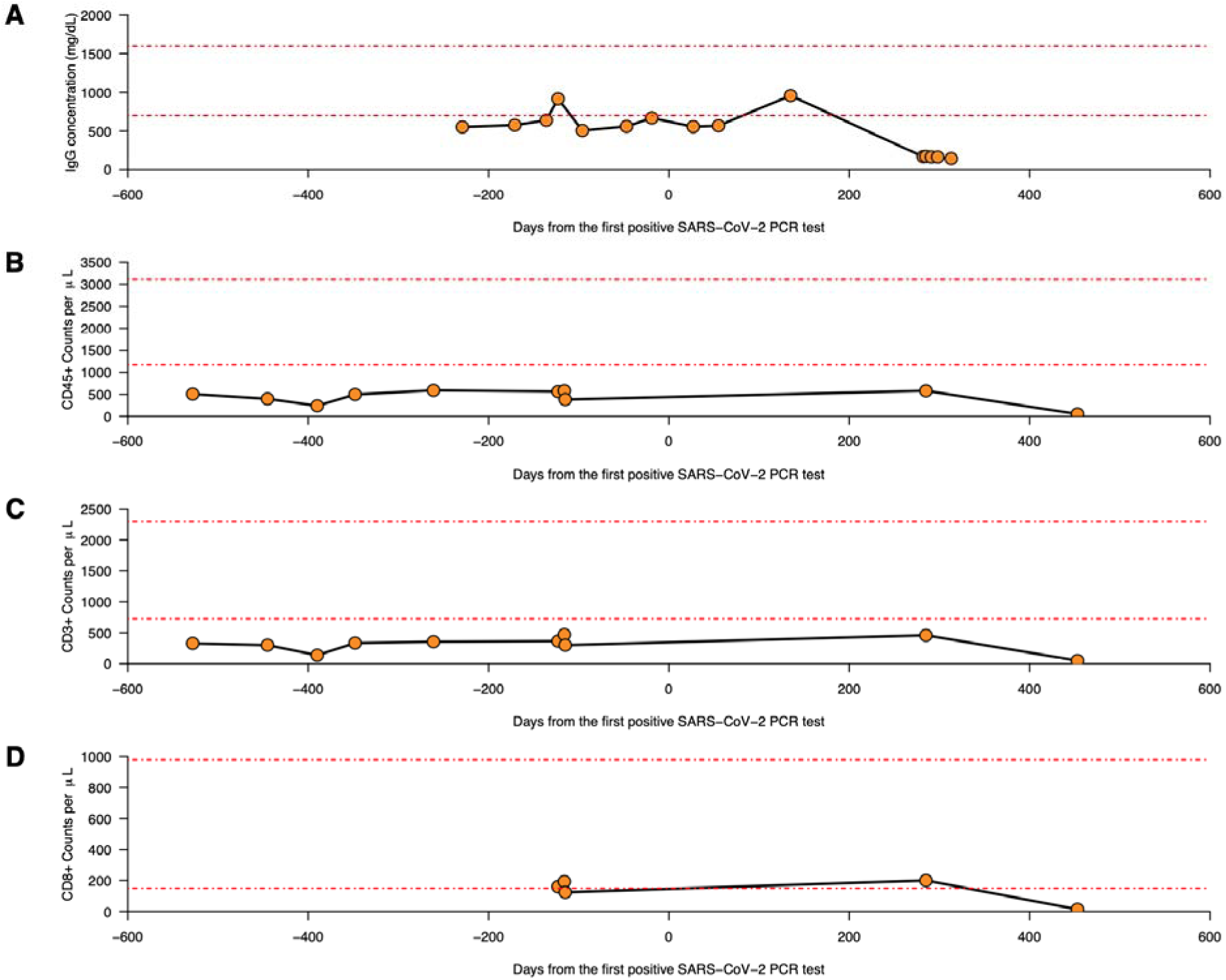
Key adaptive immune parameters as obtained from longitudinal chart reviews. (A) Serum concentration of Immunoglobulin G (IgG) (reference range from 700-1600 mg/dL are shown by the red dotted lines). **(B)** Cell counts per μl blood for CD45^+^ lymphocytes (reference range from 1170-3110/μl are shown by the red dotted lines) **(C)**, CD3^+^ T cells (reference range from 725-2300/μl are shown by the red dotted lines) and **(D)** CD8^+^ cytotoxic T cells (reference range from 150-980/μl are shown by the red dotted lines).

**Figure S2:**
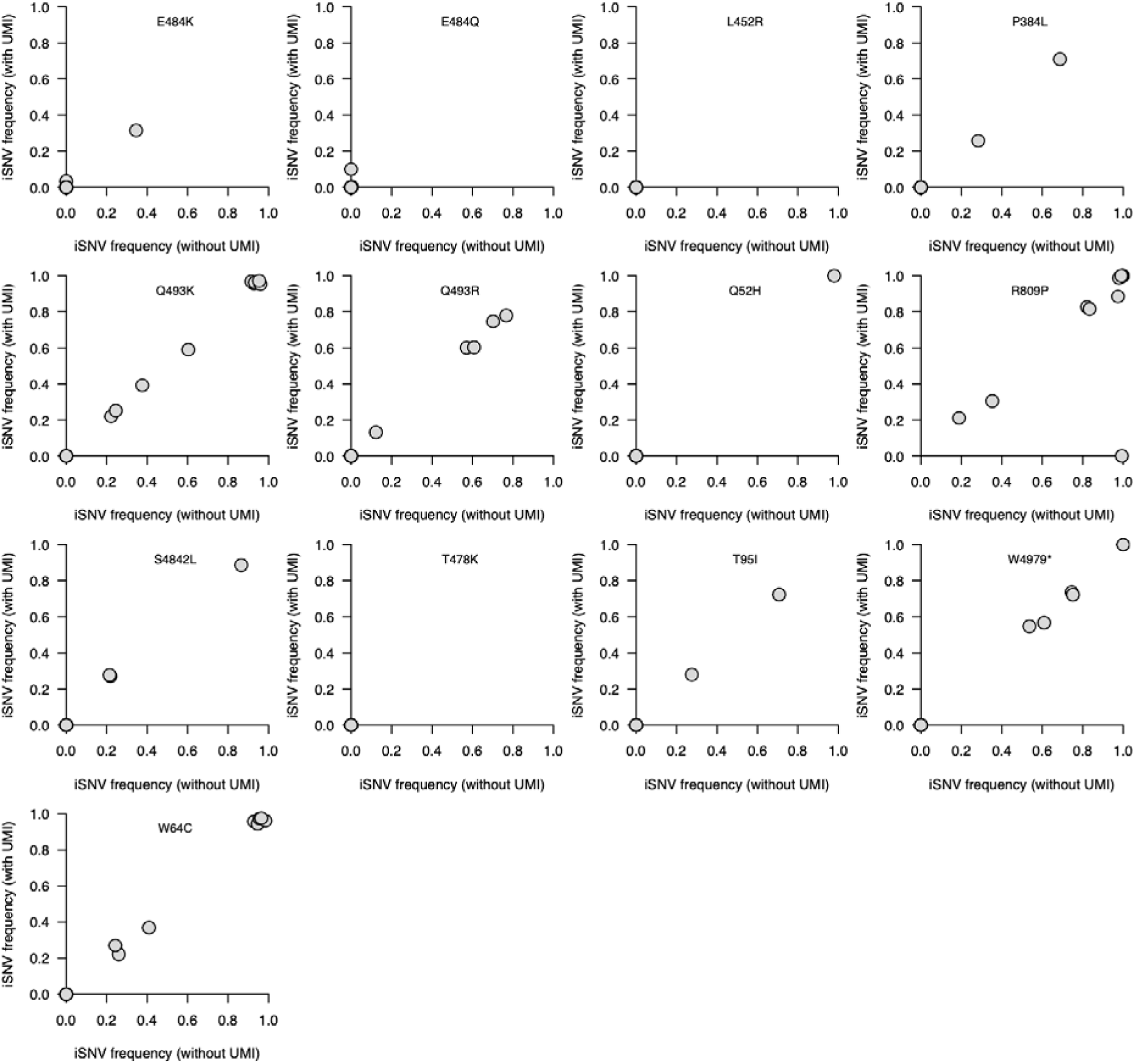
High concordance of the iSNV or mutation frequency of the spike glycoprotein mutations between samples deep-sequenced with and without unique molecular identifiers (UMI). The genomic data without the UMIs were deep-sequenced using routine amplicon-based sequencing protocol using ARTIC V3, V4, and V4.1 primers for amplicon generation using 2×150 bp paired-end reads on an Illumina NovaSeq while the samples deep-sequenced using MiSeq 2×300 bp paired-end sequencing using the Primer ID next generation sequencing protocol.

**Figure S3:**
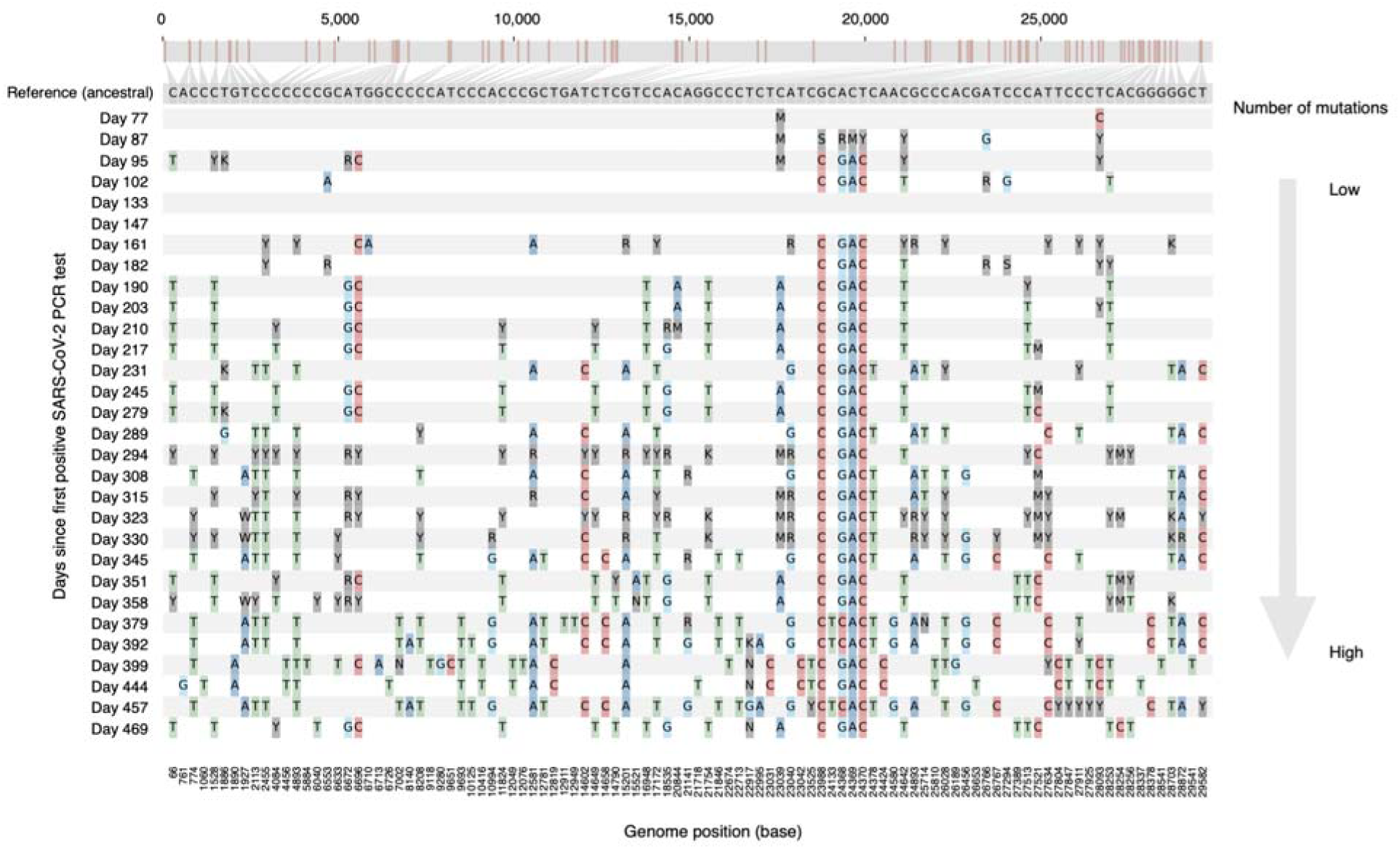
Nucleotide variation in B.1.517 SARS-CoV-2 genomes longitudinally sampled from a chronically infected patient. Consensus sequences for each genome were compared against a reference genome, a reconstructed ancestral sequence for all the chronic infection genomes.

**Figure S4:**
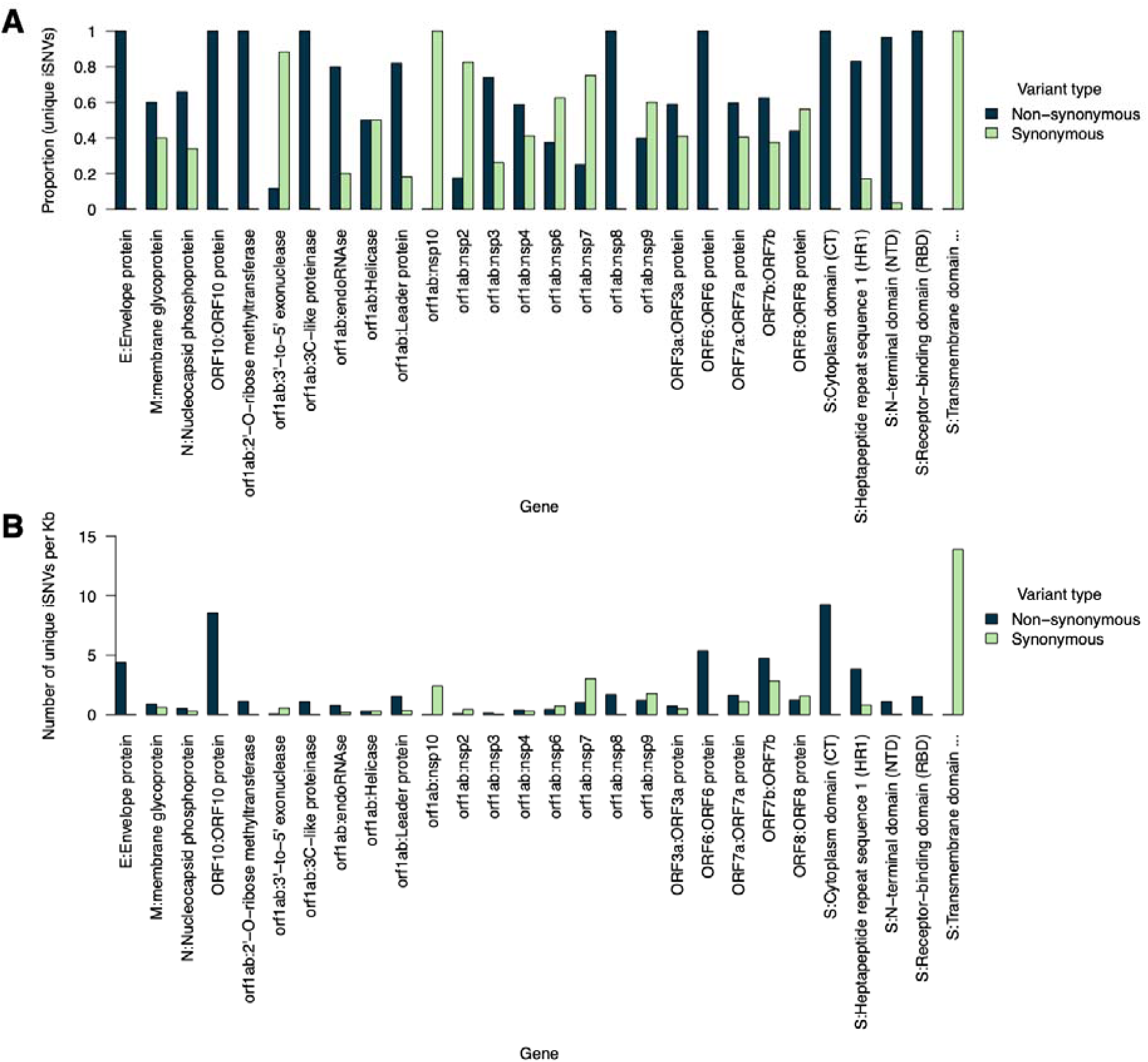
Distribution of iSNVs or mutations detected in the deep-sequenced longitudinal chronic infection samples. The y-axis shows the number of iSNVs per kilobase for different SARS-CoV-2 genomic features based on the sequence annotations in the Wuhan-1 reference genome (GenBank accession: NC_045512.2). The bars in the graph are coloured by the variant or mutation type. Additional information is provided in Data S2.

**Figure S5:**
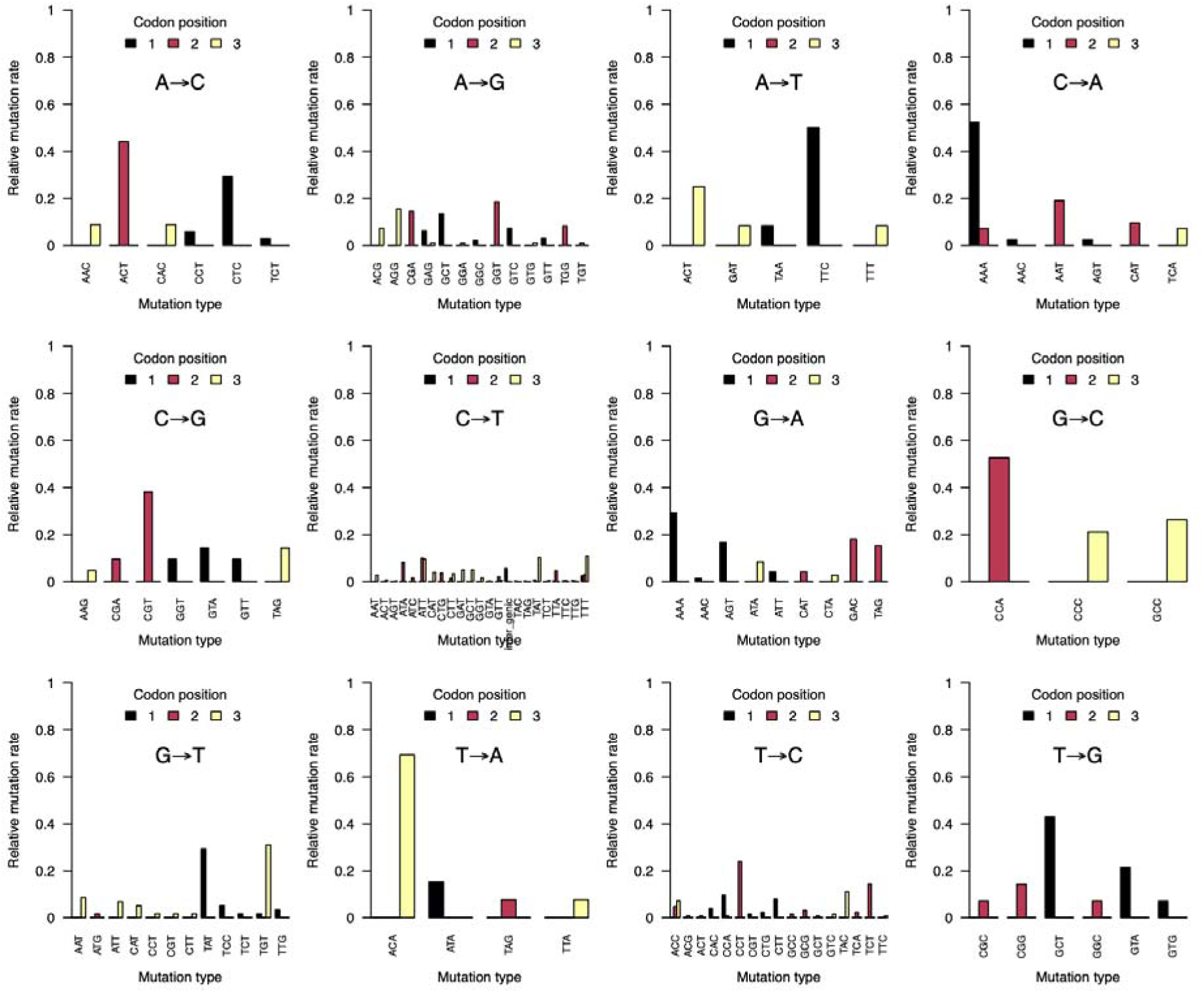
Mutation spectra of identified twelve trinucleotides during the B.1.517 chronic infection stratified by codon position. Additional information is provided provided in **Data S1**.

**Figure S6:**
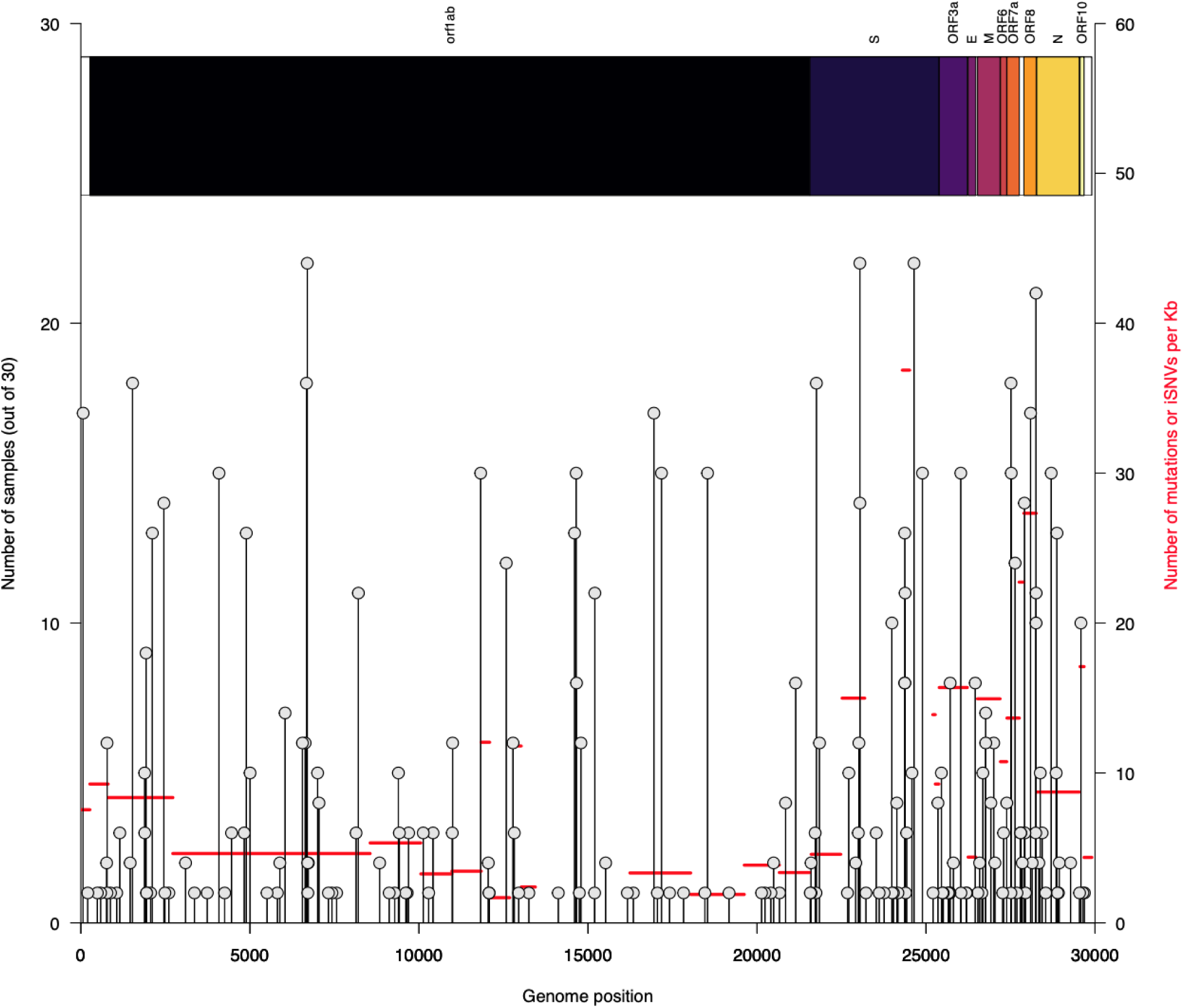
Distribution of iSNVs or mutations detected in the deep-sequenced longitudinal chronic infection samples. Graph showing the number of samples containing each unique iSNV and its position in the Wuhan-1 SARS-CoV-2 reference genome. The y-axis labels on the right side of the plot show the number of iSNVs per gene and their position in the SARS-CoV-2 genomes stratified based on the sequence feature annotations in the Wuhan-1 reference genome (GenBank accession: NC_045512.2). Additional information is provided in **Data S1**.

**Figure S7:**
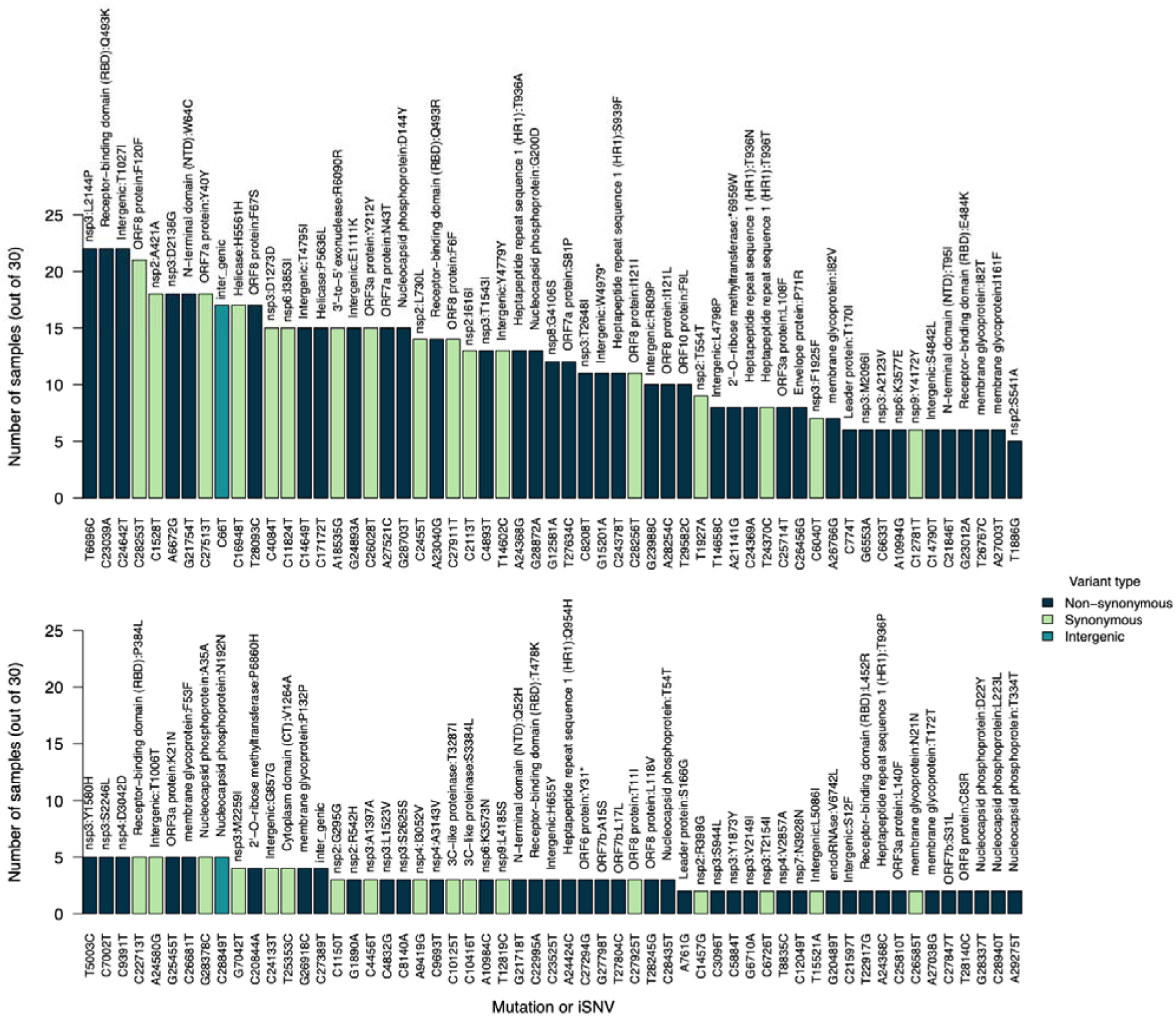
Distribution of iSNVs or mutations detected in the deep-sequenced longitudinal chronic infection samples. The x-axis labels represent iSNVs corresponding to specific nucleotide substitutions and position in the genome. The labels above the bars show the specific amino acid changes and their specific position in the SARS-CoV-2 genomes stratified based on the sequence feature annotations in the Wuhan-1 reference genome (GenBank accession: NC_045512.2). All the iSNVs are coloured by the variant or mutation type. Additional information is provided in **Data S1**.

**Figure S8:**
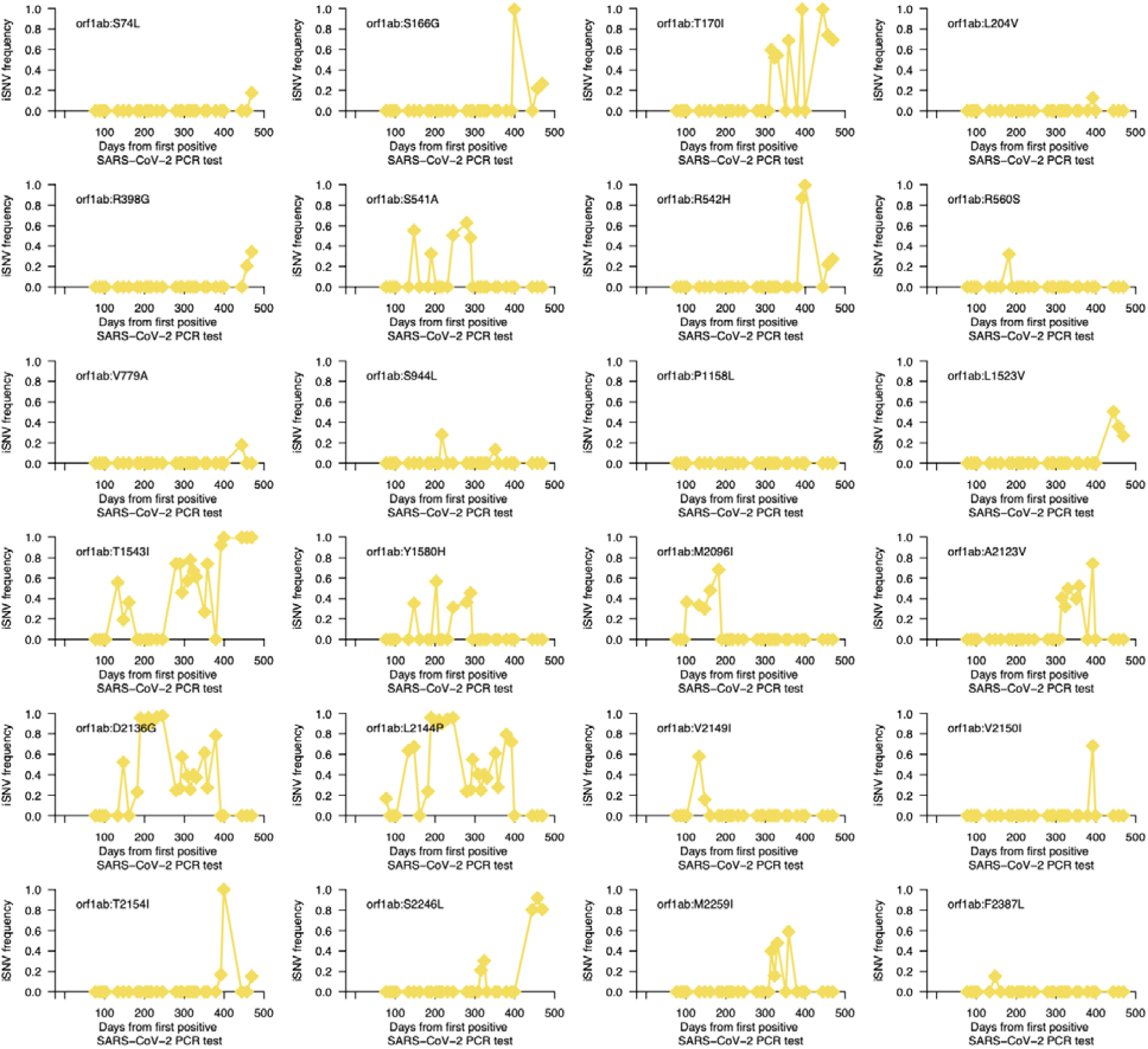
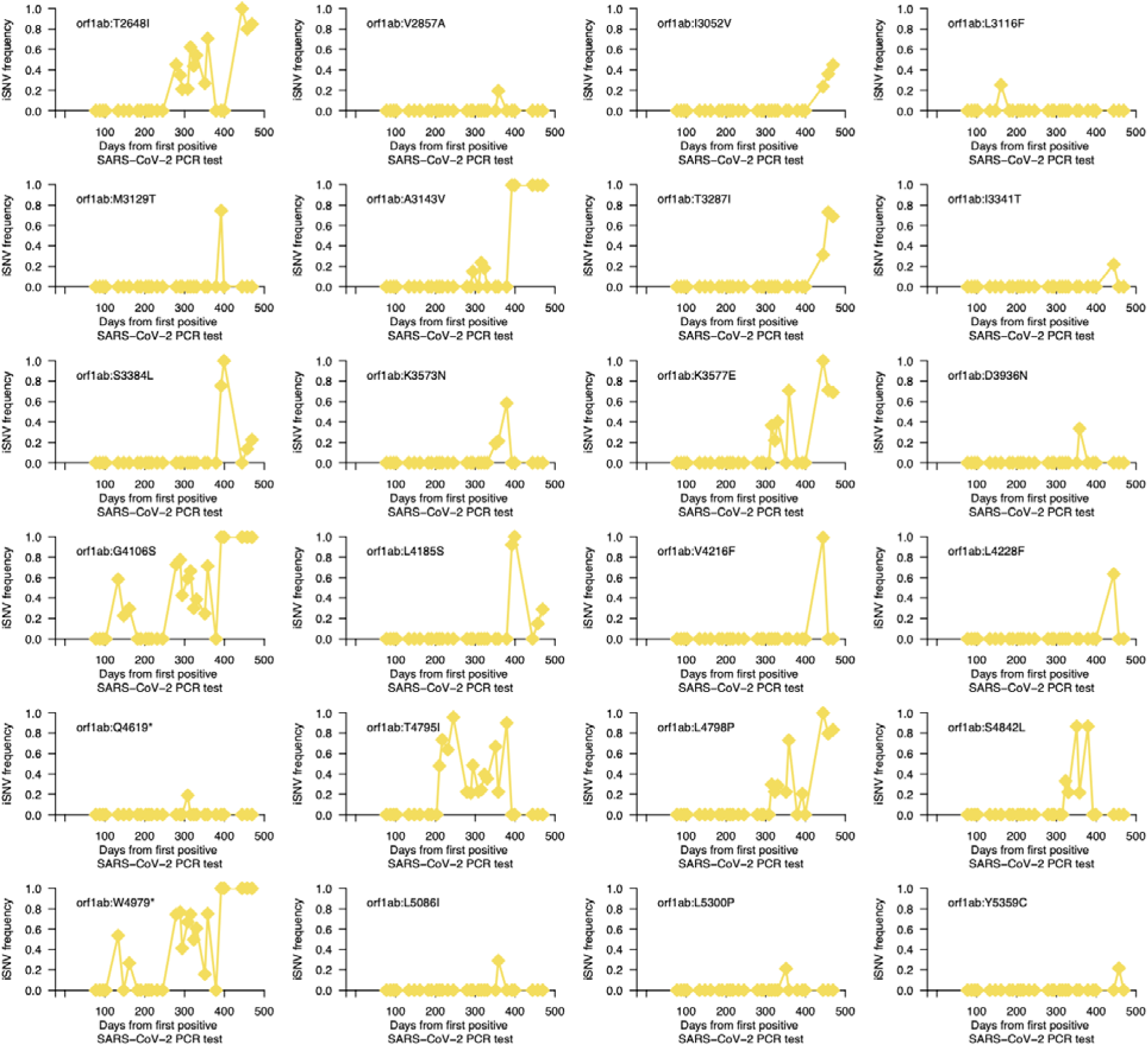
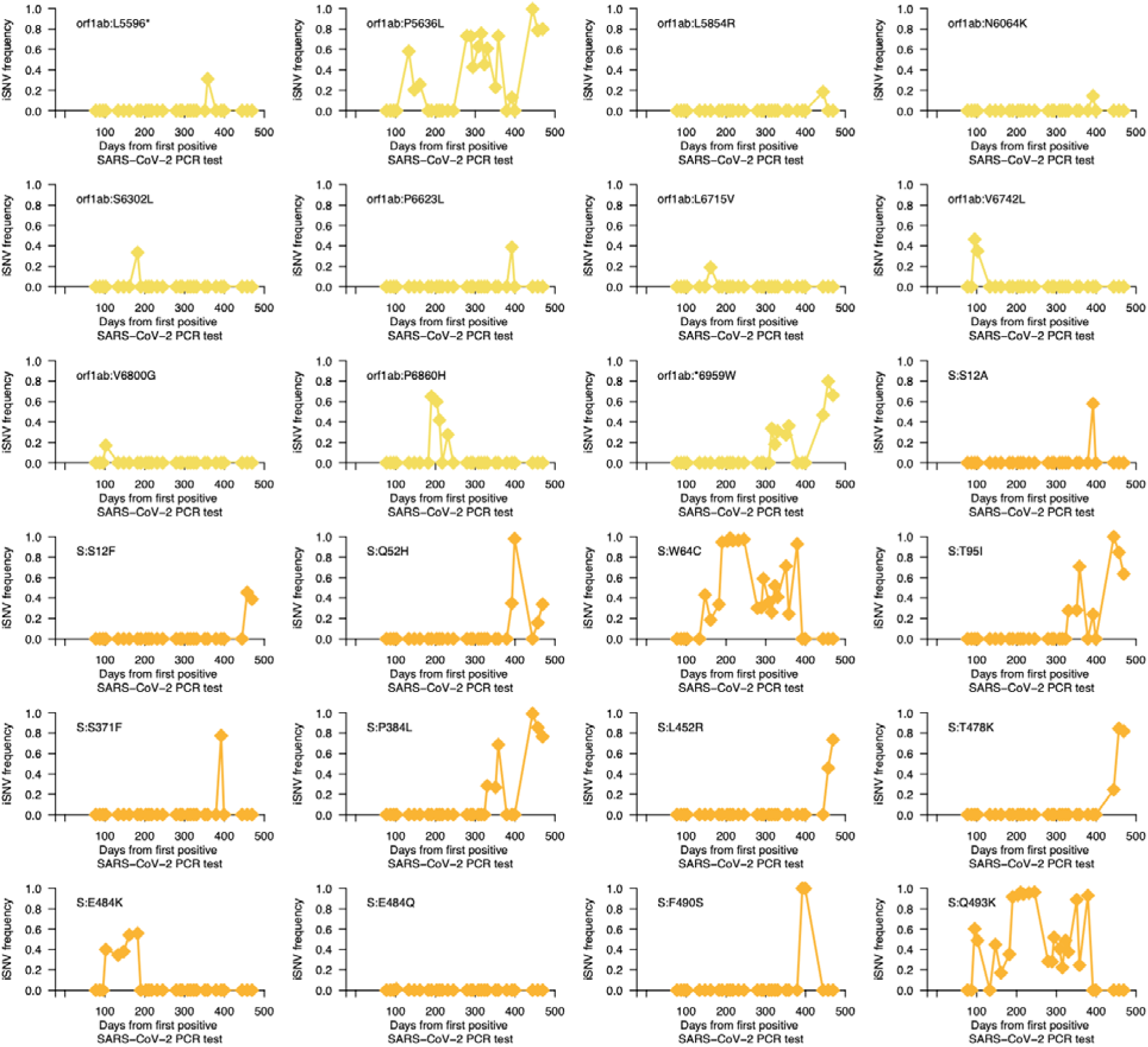
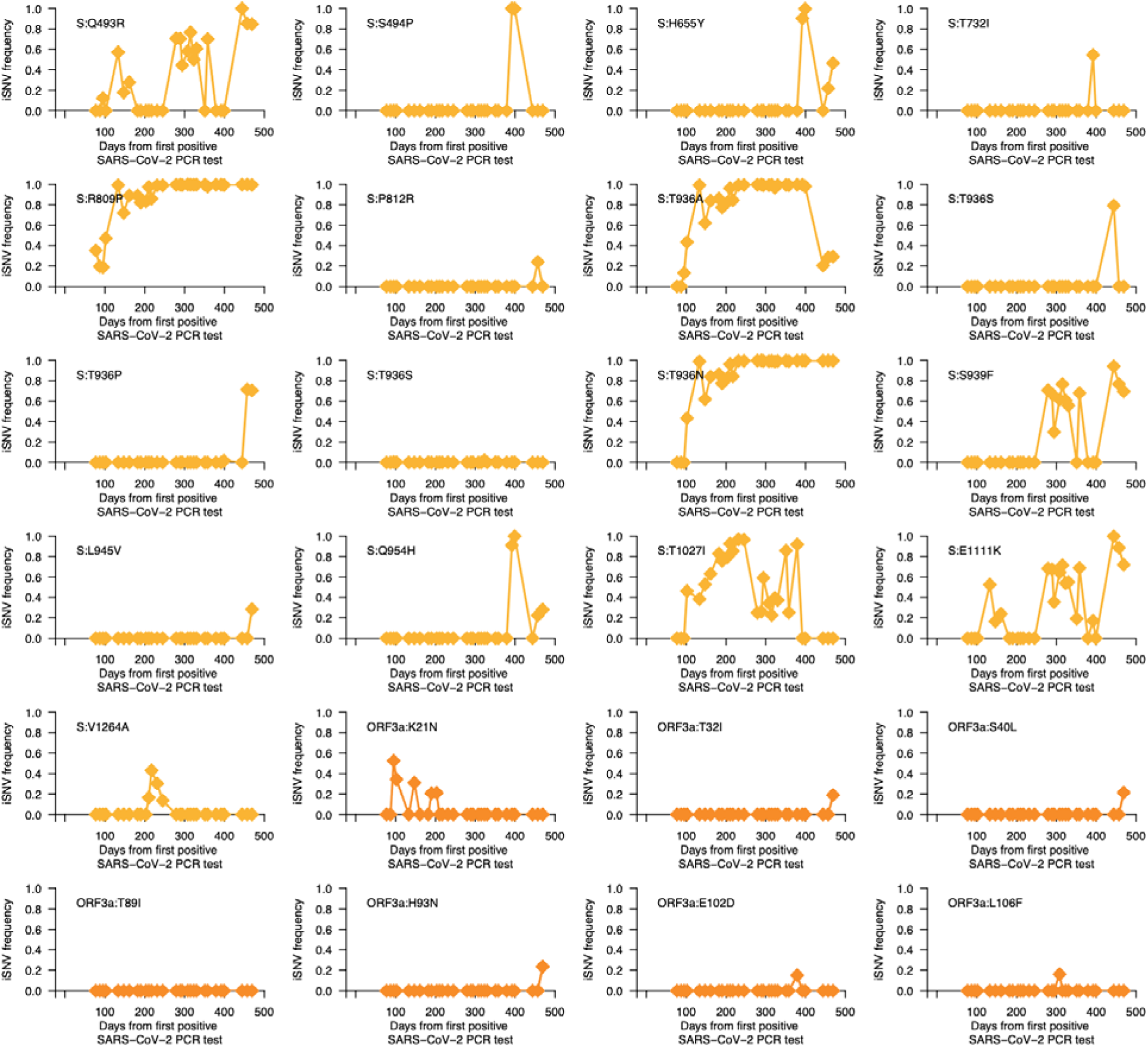
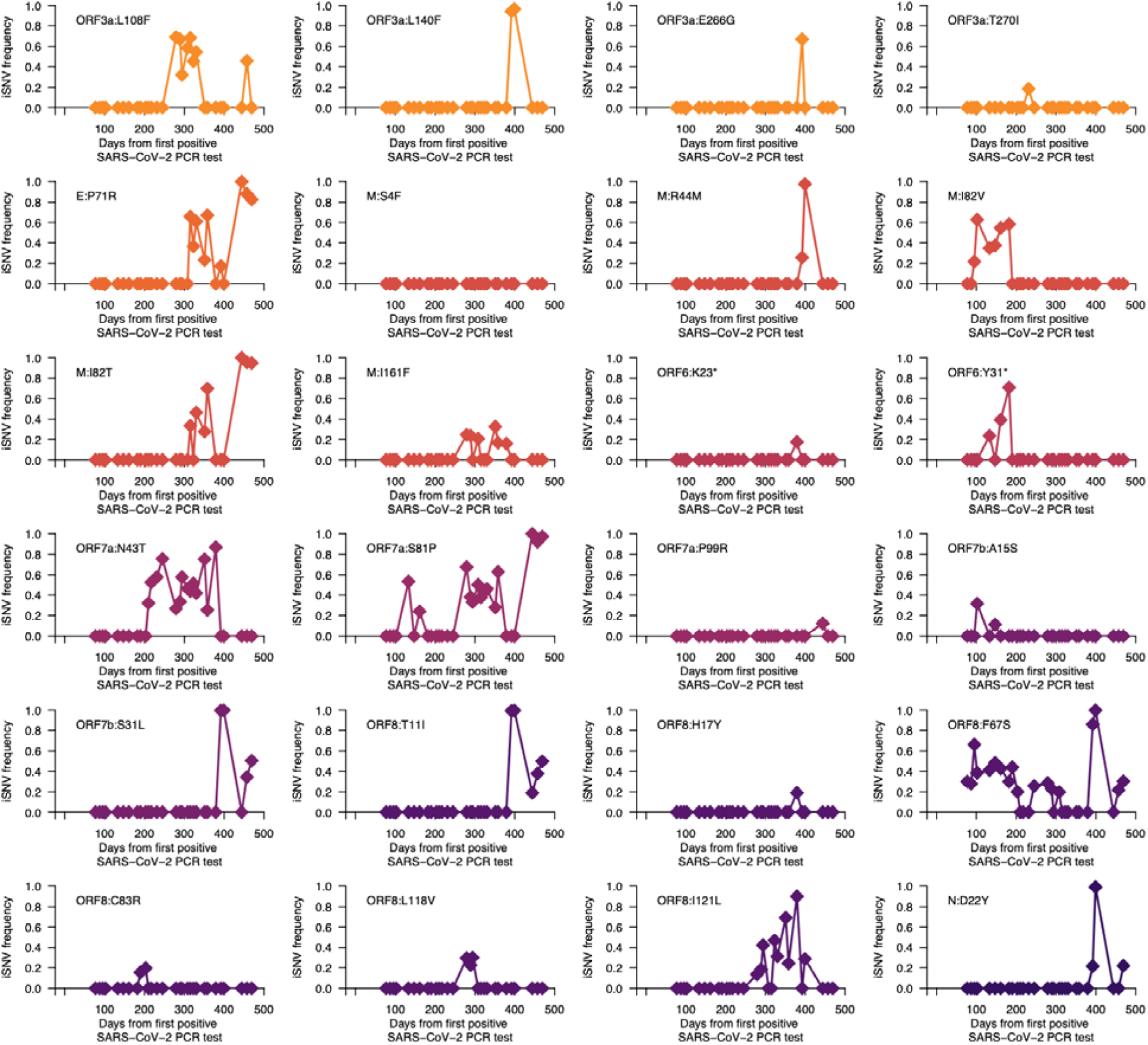
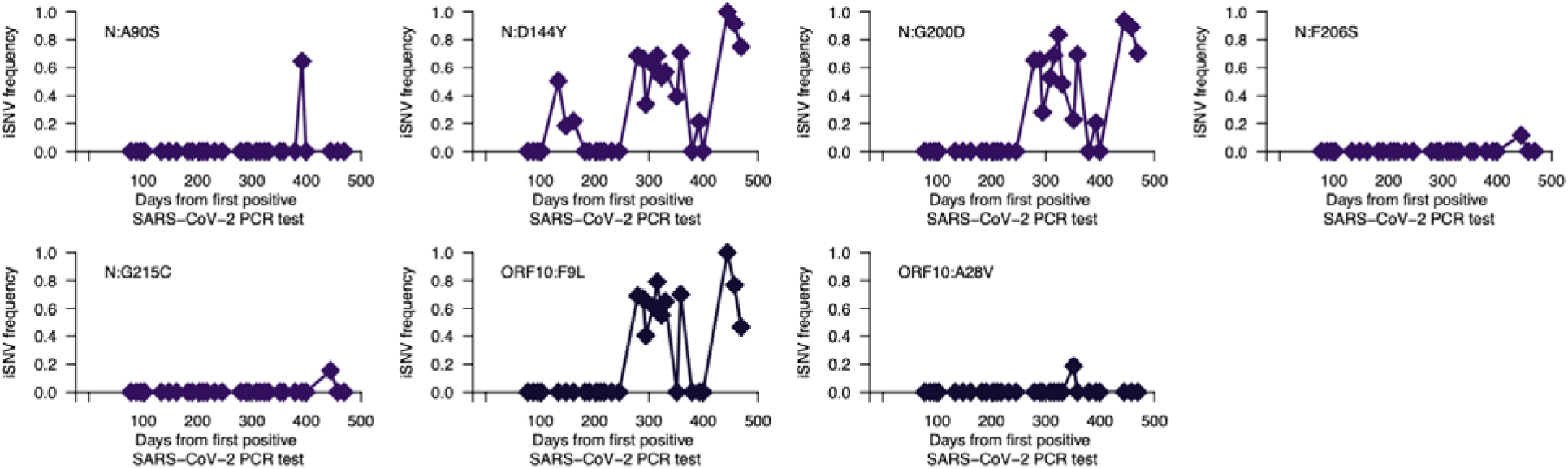
Intrahost non-synonymous mutation dynamics during chronic B.1.517 SARS-CoV-2 infection. The graphs show temporal frequencies of non-synonymous iSNVs or mutations identified in the entire genome. Additional information for all the identified mutations (intergenic, synonymous, and non-synonymous) are provided in **Data S1**.

**Table S1.** **Summary of chronic SARS-CoV-2 lineage B.1.517 infection samples included in this study.**

| Sequence ID | GISAID accession | Days since first positive RT-PCR test | RT-PCR Ct value | Virus copies per mL | Infectious virus | Primer ID sequencing | Dominant intrahost genotype | Primers |
| --- | --- | --- | --- | --- | --- | --- | --- | --- |
| hCoV-19/USA/CT-Yale-12056/2021 | EPI_ISL_10548915 | 79 | 20.5 | $3.01 \times 10^{08}$ | Yes | Yes | 1 | NEB V3 primers |
| hCoV-19/USA/CT-Yale-12057/2021 | EPI_ISL_10548916 | 89 | 22.1 | $1.13 \times 10^{08}$ | Not tested | Not done | 1 | NEB V3 primers |
| hCoV-19/USA/CT-Yale-12058/2021 | EPI_ISL_10548917 | 97 | 15.6 | $6.04 \times 10^{09}$ | Yes | Yes | 1 | NEB V3 primers |
| hCoV-19/USA/CT-Yale-12059/2021 | EPI_ISL_10548918 | 104 | 25.2 | $1.69 \times 10^{07}$ | No tested | Not done | 1 | NEB V3 primers |
| hCoV-19/USA/CT-Yale-12060/2021 | EPI_ISL_10548919 | 135 | 24.4 | $2.76 \times 10^{07}$ | Not tested | Yes | 1 | NEB V3 primers |
| hCoV-19/USA/CT-Yale-4087/2021 | EPI_ISL_2035047 | 149 | 23.2 | $5.76 \times 10^{07}$ | Yes | Yes | 1 | Illumina V3 primers |
| hCoV-19/USA/CT-Yale-12061/2021 | EPI_ISL_10548920 | 162 | 25.9 | $1.10 \times 10^{07}$ | Not tested | Not done | 1 | NEB V3 primers |
| hCoV-19/USA/CT-Yale-12062/2021 | EPI_ISL_10548921 | 184 | 28.5 | $2.25 \times 10^{06}$ | Not tested | Not done | 1 | NEB V3 primers |
| hCoV-19/USA/CT-Yale-5581/2021 | EPI_ISL_2716246 | 192 | 23.5 | $4.80 \times 10^{07}$ | Not tested | Yes | 1 | Illumina V3 primers |
| hCoV-19/USA/CT-Yale-5673/2021 | EPI_ISL_2776212 | 205 | 21.5 | $1.63 \times 10^{08}$ | Yes | Yes | 1 | Illumina V3 primers |
| hCoV-19/USA/CT-Yale-5792/2021 | EPI_ISL_2860316 | 212 | 29 | $1.66 \times 10^{06}$ | Not tested | Yes | 1 | NEB V3 primers |
| hCoV-19/USA/CT-Yale-12063/2021 | EPI_ISL_10548922 | 219 | 27.2 | $4.98 \times 10^{06}$ | Not tested | Not done | 1 | NEB V3 primers |
| hCoV-19/USA/CT-Yale-6136/2021 | EPI_ISL_3133023 | 233 | 22.3 | $1.00 \times 10^{08}$ | Not tested | Yes | 1 | NEB V3 primers |
| hCoV-19/USA/CT-Yale-6819/2021 | EPI_ISL_3370176 | 247 | 20.9 | $2.35 \times 10^{08}$ | Not tested | Yes | 1 | Illumina V3 primers |
| hCoV-19/USA/CT-Yale-9391/2021 | EPI_ISL_4198270 | 281 | 17.6 | $1.77 \times 10^{09}$ | Yes | Yes | 2 | Illumina V3 primers |
| hCoV-19/USA/CT-Yale-9977/2021 | EPI_ISL_4576991 | 291 | 23.53 | $4.71 \times 10^{07}$ | Not tested | Yes | 2 | Illumina V3 primers |
| hCoV-19/USA/CT-Yale-10101R/2021 | EPI_ISL_10548912 | 296 | 22.11 | $1.12 \times 10^{08}$ | Not tested | Not done | 1 | NEB V3 primers |
| hCoV-19/USA/CT-Yale-10960/2021 | EPI_ISL_10548913 | 310 | 29.9 | $9.54 \times 10^{05}$ | Yes | Not done | 2 | NEB V3 primers |
| hCoV-19/USA/CT-Yale-11558/2021 | EPI_ISL_5395558 | 317 | 24.6 | $2.45 \times 10^{07}$ | Not tested | Yes | 2 | NEB V3 primers |
| hCoV-19/USA/CT-Yale-11887/2021 | EPI_ISL_5639913 | 325 | 29.1 | $1.56 \times 10^{06}$ | Yes | Not done | 2 | NEB V3 primers |
| hCoV-19/USA/CT-Yale-12124/2021 | EPI_ISL_5865553 | 332 | 25.4 | $1.50 \times 10^{07}$ | Not tested | Yes | 2 | NEB V3 primers |
| hCoV-19/USA/CT-Yale-13443/2021 | EPI_ISL_7361483 | 347 | 28.3 | $2.54 \times 10^{06}$ | Yes | Yes | 1 | IDT V4 primers |
| hCoV-19/USA/CT-Yale-13444/2021 | EPI_ISL_7361527 | 353 | 32.5 | $1.94 \times 10^{05}$ | Not tested | Not done | 1 | IDT V4 primers |
| hCoV-19/USA/CT-Yale-14026/2021 | EPI_ISL_7980711 | 360 | 26.3 | $8.64 \times 10^{06}$ | Not tested | Yes | 2 | IDT V4.1 primers |
| hCoV-19/USA/CT-Yale-15439/2021 | EPI_ISL_8563219 | 381 | 21.2 | $1.96 \times 10^{08}$ | Yes | Yes | 1 | IDT V4 primers |
| hCoV-19/USA/CT-Yale-15438/2021 | EPI_ISL_8563218 | 394 | 33.6 | $9.91 \times 10^{04}$ | No | Not done | 3 | IDT V4 primers |
| hCoV-19/USA/CT-Yale-15437/2021 | EPI_ISL_8563217 | 401 | 26.1 | $9.77 \times 10^{06}$ | Yes | Yes | 3 | IDT V4 primers |
| hCoV-19/USA/CT-Yale-17291/2022 | EPI_ISL_10815044 | 446 | 34.1 | $7.30 \times 10^{04}$ | Not tested | Not done | 2 | IDT V4.1 primers |
| hCoV-19/USA/CT-Yale-17881/2022 | EPI_ISL_11025821 | 459 | 30.03 | $8.81 \times 10^{05}$ | Not tested | Not done | 2 | IDT V4.1 primers |
| hCoV-19/USA/CT-Yale-18086/2022 | EPI_ISL_11503909 | 471 | 30.9 | $5.18 \times 10^{05}$ | No | Not done | 2 | IDT V4.1 primers |

**Table S2.** Nucleotide substitution or mutation rates of the chronic infection samples and other SARS-CoV-2 variants.

| Strain | Substitutions per year (s/y) | | | Substitutions per site year (s/s/y) | | | Regression $R^2$ |
| --- | --- | --- | --- | --- | --- | --- | --- |
|  | Estimate | Lower 95% CI | Upper 95% CI | Estimate | Lower 95% CI | Upper 95% CI |  |
| Chronic infection | 35.55 | 31.56 | 39.54 | $1.21 \times 10^{-03}$ | $1.07 \times 10^{-03}$ | $1.34 \times 10^{-03}$ | 0.92 |
| All lineages | 17.16 | 16.36 | 17.96 | $5.83 \times 10^{-04}$ | $5.56 \times 10^{-04}$ | $6.11 \times 10^{-04}$ | 0.41 |
| B.1.517 (other) | 16.93 | 13.46 | 20.41 | $5.76 \times 10^{-04}$ | $4.58 \times 10^{-04}$ | $6.94 \times 10^{-04}$ | 0.51 |

**Table S3.**
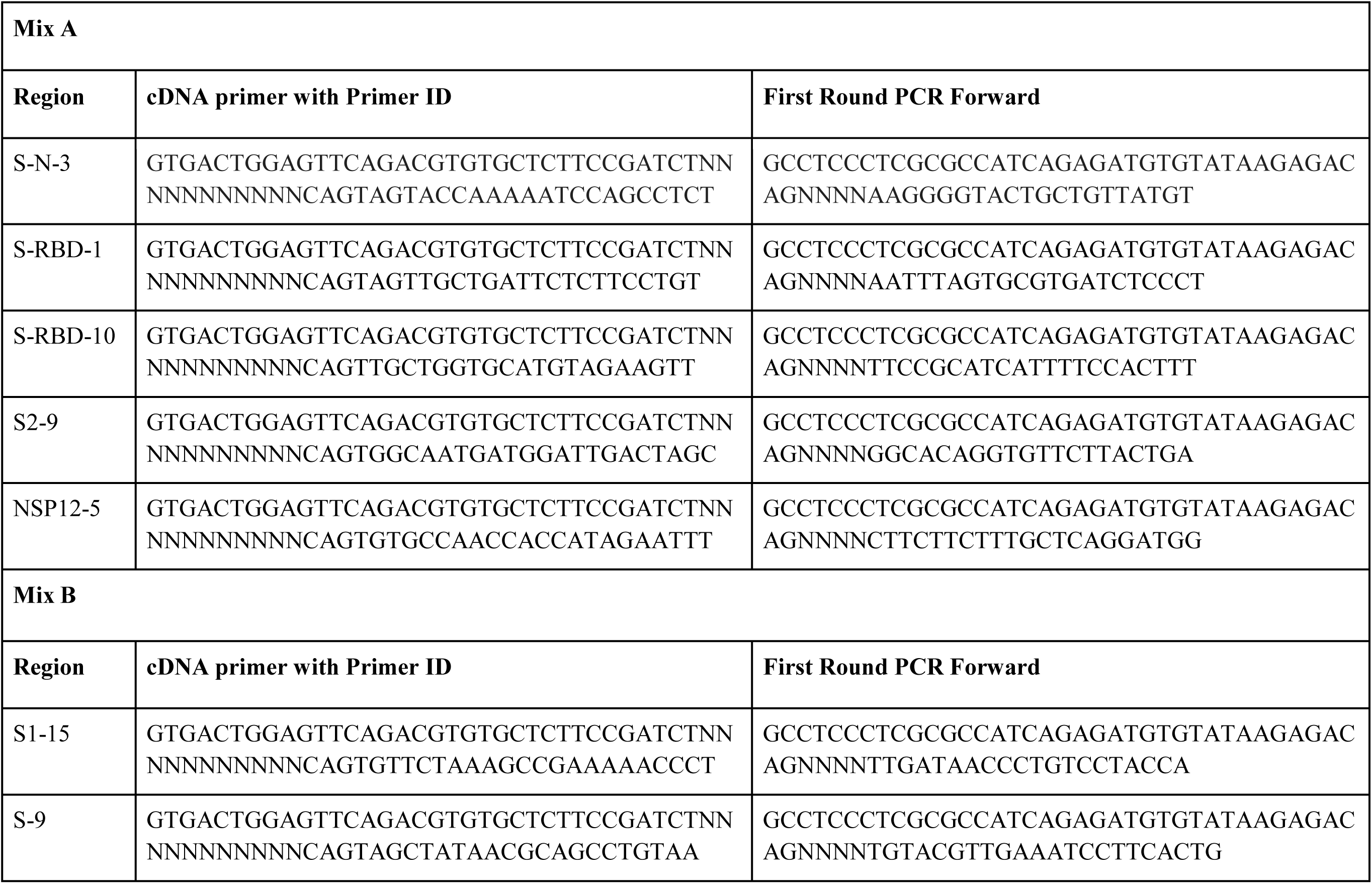
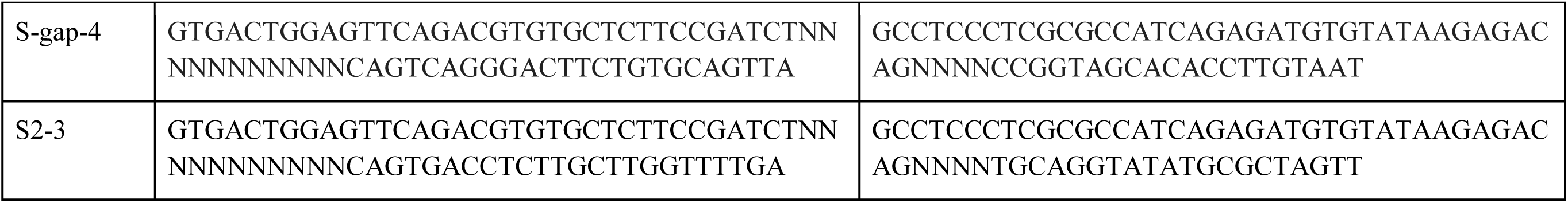
**cDNA and forward primer sequences for Multiplexed Primer ID (MPID) MiSeq library preparation for the SARS- CoV-2 S gene and nsp12.**

